# Environmental adversity moderates polygenic score effects on childhood behavioral problems in the United States

**DOI:** 10.1101/2023.06.16.23291504

**Authors:** Taylor R. Thomas, Lucas G. Casten, Jacob J. Michaelson

## Abstract

**IMPORTANCE:** Behavioral problems in children are influenced by environmental and genetic factors, but it is still unclear how much each contributes and if there are gene-by-environment interactions (GxE).

**OBJECTIVE:** Our object was to investigate how environmental adversity moderates the effects of polygenic scores (PGS) on childhood behavioral problems through additive and interaction effects.

**DESIGN, SETTING, AND PARTICIPANTS:** Participants were *N* = 7, 191 children aged 7-15 years (50% autistic) from two United States cohorts, ABCD and SPARK.

**MAIN OUTCOMES AND MEASURES:** The main outcomes were five dimensional subscales from the Child Behavior Checklist (CBCL). The genetic variables were 20 behavior-related PGS, including psychiatric diagnoses, substance use disorders, cognition, and personality PGS. Environmental adversity was estimated by the Area Deprivation Index (ADI). The ADI is a composite variable of neighborhood adversity based on education, income, and housing.

**RESULTS:** Thirteen out of the 20 PGS were significantly associated with the ADI. PGS for psychiatric and substance use disorders were positively associated with the ADI, and PGS for educational attainment and cognitive performance were negatively associated. The ADI had significant SNP heritability: *h*^2^ = 0.33 [0.24, 0.42], with the estimate similar between ABCD and SPARK. The ADI was positively associated with more behavioral problems and explained more variance than any PGS, but this effect was reduced after accounting for these potential genetic confounders. Several GxE effects were identified, including: 1.) the positive associations of the cannabis and alcohol dependency PGS with externalizing problems increased as the ADI increased, 2.) the positive associations of the anorexia PGS with thought and internalizing problems increased as the ADI increased, 3.) the positive associations of the autism PGS with internalizing problems decreased as the ADI increased, 4.) the negative associations of the educational attainment and cognitive performance PGS with several behavioral problems increased as the ADI increased, and 5.) the extraversion PGS association with social problems was negative in an advantaged environment but positive in a disadvantaged environment.

**CONCLUSIONS AND RELEVANCE:** Environmental adversity estimated by the ADI moderates the effects of some PGS on childhood behavioral problems through additive and interaction effects. This highlights the importance of considering both genetic and environmental factors in understanding childhood behavioral problems. Our findings emphasize the need to include PGS of personality and cognitive traits, in addition to psychiatric PGS.

## 1 Introduction

Adversity can have profound effects on many outcomes in childhood, including behavioral problems. Environmental adversity is typically estimated using indicators of experiences or location quality. One estimate of location adversity in the United States is the Area Deprivation Index (ADI) [1]. The ADI percentile ranks every neighborhood based on education, income, and housing characteristics. Each neighborhood across the United States has an ADI percentile, with 1 for the most advantaged neighborhoods and 100 for the most disadvantaged neighborhoods. The ADI has been associated with a range of health outcomes. For example, in a large sample of children in the United States (the Adolescent Brain Cognitive Development study (ABCD) [2]), a higher ADI was associated with worse performance on cognitive tests [3] and lower hippocampal volumes [4].

In addition to adversity, behavioral problems and psychiatric conditions in childhood are also affected by genetic factors [5, 6]. However, the effects of environmental versus genetic factors are difficult to disentangle, in part due to the environmental variables themselves having genetic associations (i.e., gene-environment correlates). For example, adverse childhood experiences like maltreatment, parental criminality, and parental separation are associated with increased behavioral problems, but these adverse childhood experiences are also associated with increased genetic risk for psychiatric conditions [7]. As another example, for adults in the UK, both birthplace and current address were strongly associated with educational attainment and income polygenic scores (PGS) [8]. Controlling for these gene-environment correlates attenuated the genetic effects of many traits, most prominently BMI and substance use.

Despite the potential genetic confounding of environmental variables, recent gene-by-environment interaction (GxE) studies have made headway in elucidating how the environment modulates the effects of genetic factors on psychiatric disorders and behavioral problems. For example, genome-by-trauma interactions explained a significant proportion of depression variability in UK Biobank participants [9]. In another UK study, SNP-by-environment genome-wide association studies (GWAS) identified several SNPs that had significant interactions with the Townsend Index (which is similar to the ADI) for psychiatric disorders [10]. A study of physicians in the United States found that the depression PGS was most predictive of depressive symptoms for physicians when they were under stress during their first year of residency [11]. These GxE effects estimate the penetrance of the genetic effect. Penetrance is defined as the extent to which a genetic effect is expressed at different levels of the environment. In other words, penetrance describes how the environment suppresses or amplifies a genetic effect [12].

The majority of these GxE studies have used European adult cohorts. European countries have lower income inequality and poverty rates than the United States [13, 14]. Therefore, studies in the United States may be better positioned to detect GxE effects. In this study, our aim was to investigate how the ADI moderates the effects of 20 PGS on childhood behavioral problems in the United States. We combined datasets from two nationwide studies: ABCD [2] and SPARK [15]. The ABCD cohort was not recruited based on the presence or absence of any particular diagnosis and therefore the variability in behavioral problems was low. In order to expand the variance in behavioral problems, we combined ABCD with the SPARK study [15], which is an autism cohort.

We first tested for gene-environment correlates of the ADI by performing associations of 20 PGS and calculating the SNP heritability of the ADI. Next, we performed head-to-head comparisons of the ADI versus PGS to investigate which risk factor had stronger independent effects on childhood behavioral problems. Then, we tested for additive and interaction effects between the ADI and PGS. Lastly, we further investigated these interaction effects by calculating the average marginal effects of PGS across the range of the ADI, which effectively captures how the PGS penetrance changes with respect to varying levels of environmental disadvantage.

## 2 Materials and methods

### 2.1 Childhood behavioral problem variables: Child Behavior Checklist subscales

The estimates of childhood behavioral problems were from the Child Behavior Checklist (CBCL) [16]. The CBCL is a parent-report questionnaire of 118 Likert-scale (0, 1, 2) items that cover a broad range of behavioral and emotional problems. The CBCL is suitablea for children ages 6 to 17. Specific CBCL items can be summed to form various quantitative domain subscales. We used the five syndromic subscales: attention problems, social problems, thought problems, internalizing problems (anxious, withdrawn depressed, and somatic complaints), and externalizing problems (aggressive and rule-breaking behavior). For our analyses, the raw summed scores were accounted for age in months by quasi-Poisson regression residualization and then Z-scaled (i.e., centered to a mean (*µ*) of 0 and a standard deviation (*σ*) of 1).

### 2.2 Environmental adversity variable: Area Deprivation Index

The estimate of environmental adversity was the Area Deprivation Index (ADI) [1] national percentiles. The ADI is a composite variable of disadvantage from neighborhood characteristics related to education, income, and housing. We used the 2015 ADI because it was what was available for both ABCD and SPARK.

### 2.3 Study participants

We used participants from two nationwide cohorts in the United States. The first cohort, the Adolescent Brain Cognitive Development (ABCD) study [2] is a cohort that is representative of the general population (i.e., was not recruited based on the presence or absence of any particular diagnosis). The second cohort, the SPARK study [15], is an autism cohort. Both cohorts were recruited from various locations throughout the United States.

For ABCD, we used Data Release 4. The ABCD study collected the CBCL at intake and then each year thereafter for three years. Therefore, the CBCL ages at baseline ranged from approximately 8 to 10 years and at three-year follow-up ranged from 12 to 14 years. Because SPARK had a higher range in CBCL ages, we randomly sampled one CBCL for each ABCD participant. No CBCLs for ABCD participants had missing data. For the ADI, we used the 2015 ADI associated with the participant’s first address in the residential history data field. This is presumably the address at the time of study enrollment.

For SPARK, the Version 9 phenotypic data release was used. Version 9 contains the ADI based on the participant’s address (again presumably the address at the time of study enrollment). In order to be consistent with the ABCD ADI, we removed any participants that reported the 2020 ADI and only included the 2015 ADI. The majority of the CBCLs were also from Version 9, while a subset were from a SPARK Research Match [17] (the Research Match study was approved by the University of Iowa IRB (IRB 202002251) and is described in detail in [17]). We removed participants with more than 9 missing items from the CBCL. This left us with 0.19% missing CBCL data. We imputed these missing data using predictive mean matching using the MICE algorithm and R package [18].

After these initial phenotype gathering steps, we had CBCLs for *N* = 19, 879 participants from ABCD (*N* = 9, 839) and SPARK (*N* = 10, 040). We then retained only those with the 2015 ADI available (*N* = 19, 322 remaining). Next, we removed those in SPARK who reported a known, diagnosed genetic syndrome (*N* = 19, 100 remaining). Next, we only kept participants if they had genetic data that passed our quality control and were in the majority genetic ancestry based on genetic principal components clustering (see next section); this left us with *N* = 10, 626 participants. Next, we only kept participants with their CBCL age between 7 and 15 (*N* = 9, 830 remaining). We removed participants that were extreme outliers (above the 99.5^th^ percentile in our sample, which was a score of 7). Recent work by [19] found low CBCL scores to be unreliable in ABCD. Therefore, we were strict with our inclusion criteria for low total scores and removed participants that were below the 20^th^ percentile (a score of 7). This left us with *N* = 7, 962. Lastly, we then filtered participants based on a relatedness threshold of 0.05 using GCTA [20]. This left us with our final sample size of *N* = 7, 191.

### 2.4 Genotype quality control

The methods for genotype quality control, imputation, and calculation of genetic principal components and polygenic scores have been previously described in [21]. The ABCD genotypes were quality controlled for missingness and contamination before release, so no additional quality control was performed prior to genotype imputation. For SPARK, we used the genotypes from the integrated whole-exome-sequencing (iWES1) 2022 Release and the SPARK whole-genome-sequencing (WGS) Release 2, 3, and 4. SPARK iWES1 (*N* = 69, 592) was quality controlled on release, including removing samples due to heterozygosity or high missingness, so no further quality control was performed by us before genotype imputation. SPARK iWES1 also provided genetic ancestry assignments based on the 1000 Genomes populations [22]. SPARK WGS Release 2 (*N* = 2, 365), Release 3 (*N* = 2, 871), and Release 4 (*N* = 3, 684) were not quality controlled on release, so we performed quality control using PLINK [23] before genotype imputation. First, we removed participants from the WGS releases if they were in iWES1. Second, we removed variants with missingness higher than 0.1 and participants with missingness higher than 0.2. Third, we merged the three releases and then removed any participant whose heterozygosity (F statistic) was not within 3 standard deviations of the mean heterozygosity across the three releases. We then used the TopMed reference panel [24] to identify strand flips. The final sample size for WGS 2-4 was *N* = 8, 152.

### 2.5 Genotype imputation and merging

ABCD, SPARK iWES1, and SPARK WGS 2-4 (quality controlled) were imputed to the TopMed [24] reference panel using the Michigan Imputation Server [25] with the phasing and quality control steps included and to output variants with imputation quality r2 *>* 0.3. After imputation, the variants were filtered to only the HapMap SNPs (*N* = 1, 054, 330 variants) with imputation quality r2 *>* 0.8 using bcftools [26]. They were lifted over from hg38 to hg19 using the VCF-liftover tool (https://github.com/hmgu-itg/VCF-liftover) and the alleles normalized to the hg19 reference genome. Finally, the files were merged and only variants with 0% missingness were retained (*N* = 914, 328).

### 2.6 Genetic ancestry and principal components

Genetic principal components (PCs) were calculated using the bigsnpr package [27], specifically by following the author’s recommendations [28] and their tutorial: https://privefl.github.io/bigsnpr/articles/bedpca.html. In summary, we 1.) used the `snp_plinkKINGQC` function to identify and remove related participants at the KING threshold of 2*^−^*^3.5^, 2.) performed principal component analysis using the `bed_autoSVD` on just the unrelated participants, 3.) detected principal component outliers and removed them, 4.) recalculated the principal components, and 5.) projected the principal components onto the entire cohort using the `bed_projectSelfPCA` function. We assigned genetic ancestry by performing k-means clustering with the top 40 principal components with K = 5 (for the five populations from 1000 Genomes [22]) and used the genetic ancestry labels from iWES1 to assign labels to the five genetic population clusters. We then only used participants that filtered in the majority European cluster.

### 2.7 Polygenic score calculations

Polygenic scores (PGS) were calculated using LDpred2 [29] and the bigsnpr tools [27] in R [30]. Because SPARK is family-based, an external LD reference based on 362,320 individuals in UK Biobank (provided by the authors of LDpred2) was used to calculate the genetic correlation matrix, estimate heritability, and calculate the infinitesimal beta weights. PGS were calculated from the following individual genome-wide association studies: ADHD [31], autism [32], major depressive disorder (MDD) [33], bipolar disorder [34], schizophrenia (SCZ) [35]. anorexia [36], insomnia [37], alcohol dependency [38], cannabis use disorder [39], and educational attainment (EA) [40]. The PGS for the five cognitive traits are all from the same study [41]: general cognitive performance (cog gen or gFactor), executive functioning (EF, tower rearranging test), non-verbal reasoning (NVR, matrix pattern recognition test), working memory (WM, memory pairs-matching test), and reaction time (RT). The public LDpred2 beta weights from the Polygenic Index Repository [42] were used to calculate PGS for extraversion [43], neuroticism [44], openness [45], nicotine dependency (cigarettes per day) [46], and BMI [47].

We filtered the PGS to only our final sample (*N* = 7, 191) and then accounted for the 10 genetic principal components by linear regression residualization. The PGS were then Z-scaled to a mean (*µ*) of 0 and a standard deviation (*σ*) of 1.

### 2.8 SNP-based heritabilities

SNP-based heritabilities were calculated using GCTA [20], specifically GCTA REML [48]. The 10 genetic PCs were used as covariates in GCTA.

### 2.9 Statistical analyses

The analyses were performed in R [30]. All variables were Z-scaled prior to model input (mean (*µ*) = 0 and standard deviation (*σ*) = 1). Effects are reported with the 95% confidence intervals in brackets. Multiple testing correction was performed for the 20 PGS and five CBCL phenotypes with the FDR method (Benjamini and Hochberg [49]).

The Breusch-Pagan (BP) test for heteroscedasticity [50] was performed using the bptest function from the lmtest R package [51]. The Breusch-Pagan test examines whether there is a significant relationship between the squared residuals and the independent variable by augmenting the regression model. The coefficient of the squared residuals in the augmented model, which is the BP test statistic, measures the extent to which the variance of the residuals depends on the values of the independent variable(s).

Average marginal effects (AME) were calculated using the sim_slopes function from the interactions R package [52]. AME were calculated along the range of the ADI at every 0.5 standard deviations from the mean ADI (*µ* = 0 and *σ* = 1). The minimum ADI was -2.4 and the maximum was 3.6, so we calculated the AME at ADI -2.0, -1.5, -1.0, -0.5, 0, 0.5, 1.0, 1.5, 2.0, 2.5, 3.0.

## 3 Results

### 3.1 Area Deprivation Index genetic signals

#### 3.1.1 Participant demographics

The participants were *N* = 7, 191 unrelated children from two United States nationwide cohorts: *N* = 3, 719 from ABCD, a sample representative of the general population, and *N* = 3, 472 from the SPARK autism cohort (Table 1). All of the children from SPARK were autistic, while 2% of the children from ABCD were autistic. The ages ranged from 7 to 15 years old, with an average age of *µ* = 11.3 (*σ* = 2) (Figure S1A). The sample was minority female (34%), primarily due to the sex imbalance in the SPARK sample (21% female). The ABCD sample was balanced on sex (45% female). All of the participants were of European genetic ancestry based on clustering with the genetic principal components (PCs).

**Table 1.** Participant demographics. The N and percent of total in parenthesis are reported for sex and autism diagnosis. The mean and standard deviation in parentheses are reported for age, the Child Behavior Checklist (CBCL) total score, and the Area Deprivation Index (ADI).

| variable | combined | ABCD | SPARK |
| --- | --- | --- | --- |
| sample size | 7,191 | 3,719 | 3,472 |
| sex (female) | 2,417 (34%) | 1,682 (45%) | 735 (21%) |
| autism | 3,561 (50%) | 89 (2%) | 3,472 (100%) |
| age (years) | 11.3 (2) | 11.2 (1.3) | 11.4 (2.5) |
| CBCL total | 40.8 (29) | 23.3 (16.8) | 59.6 (27.5) |
| ADI | 40.6 (24.8) | 34.1 (22.6) | 47.5 (25.1) |

#### 3.1.2 Area Deprivation Index distribution and weighted linear regression

Our environmental adversity variable was the Area Deprivation Index (ADI) 2015 national percentile. The ADI is a composite variable of neighborhood disadvantage based on education, income, employment, housing quality, and household characteristics. The ADI ranges from 1 being the lowest areas of deprivation (i.e., the most advantaged environments) and 100 being the highest areas of deprivation (i.e., the most disadvantaged environments). Figure 1A shows the map of the ADI across the United States from [53]. For our sample, we used the participant’s ADI from their address given when they enrolled in the ABCD or SPARK study. Figure 1B shows the ADI distribution in our sample. The SPARK participants had an average ADI of *µ* = 47.5 (*σ* = 25.1). The ABCD participants were on average in more advantaged environments, with an average ADI of *µ* = 34.1 (*σ* = 22.6). This difference between ABCD versus SPARK was significant: *x* = 13.5 [12.4, 14.6]*, p <* 0.001*, d* = 0.57. The known true population distribution of the ADI is uniform with a median of 50, but our sample was not uniformly distributed. Therefore, we used our sample distribution to generate weights so that each ADI percentile contributed equally to subsequent linear regression models (Figure S2).

**Figure 1.**
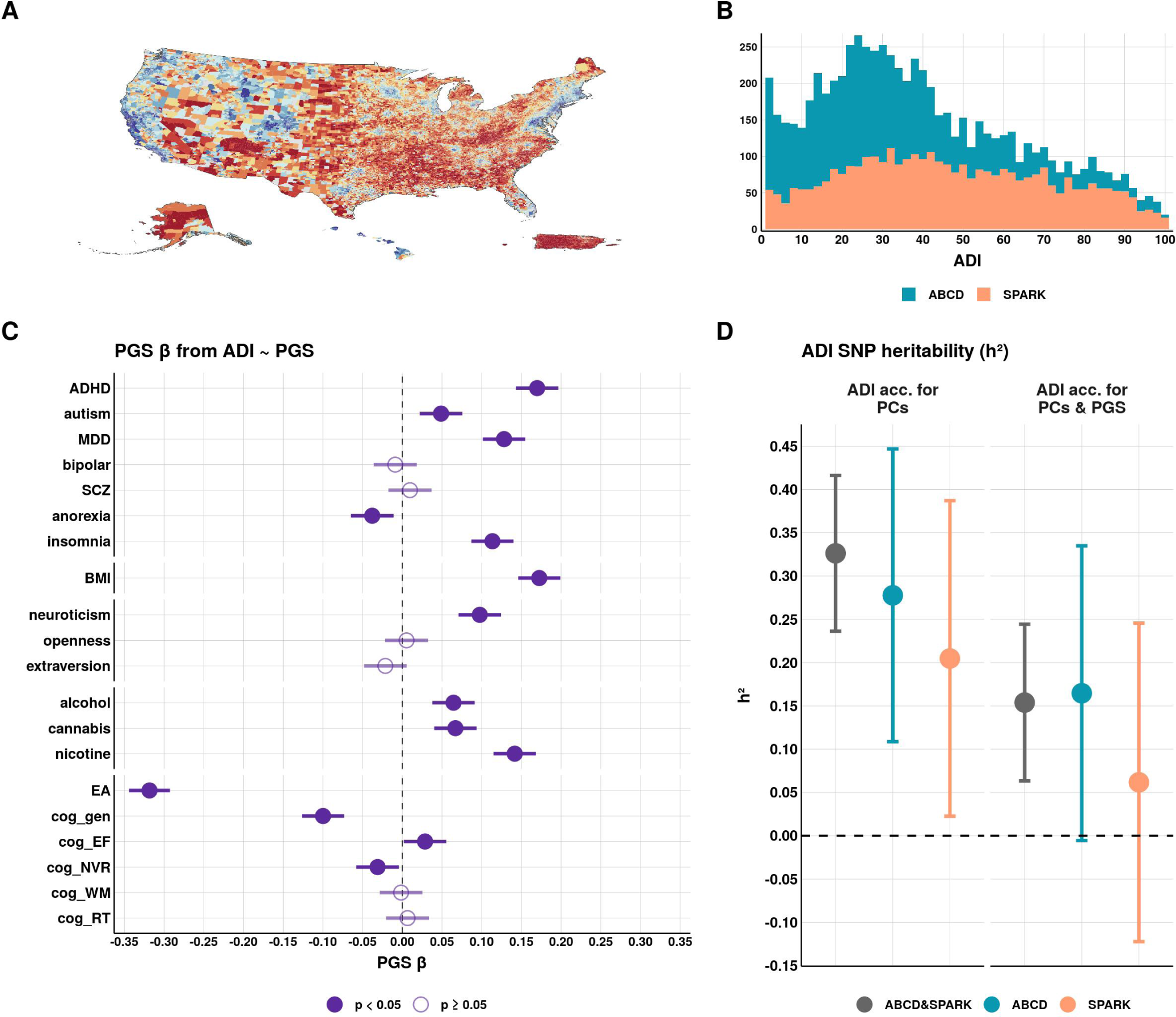
Genetic signals of the Area Deprivation Index. The environmental adversity variable in our study was the Area Deprivation Index (ADI) national percentiles **(A)** United States map of the ADI downloaded from [53], with blue indicating the most advantaged environments (ADI = 1) and red indicating the most disadvantaged environments (ADI = 100). **(B)** Distribution of the ADI in our sample of *N* = 7, 191 (ABCD *N* = 3, 719, SPARK *N* = 3, 472). **(C)** Associations of 20 polygenic scores (PGS) with the ADI. The PGS *β* estimates are with the 95% confidence intervals from the weighted linear regression model: *ADI ∼ PGS*. Prior to model input, the ADI and PGS were accounted for the 10 genetic principal components (PCs) by linear regression residualization. Associations not accounting for the 10 genetic PCs are in Table S1. The associations calculated separately for ABCD and SPARK are in Figure S4. **(D)** SNP-based heritability (*h*^2^) of the ADI before and after accounting for the 20 PGS (in addition to the 10 genetic PCs). The *h*^2^ estimates are with the 95% confidence intervals. PGS: **MDD** = major depressive disorder, **SCZ** = schizophrenia, **alcohol** = alcohol dependency, **cannabis** = cannabis use disorder, **nicotine** = nicotine dependency, **EA** = educational attainment, **cog gen** = general cognitive performance, **cog EF** = executive functioning, **cog NVR** = non-verbal reasoning, **cog WM** = working memory, **cog RT** = reaction time.

#### 3.1.3 Polygenic score associations with the Area Deprivation Index

Our genetic variables were polygenic scores (PGS) for 20 behavior-related traits: seven psychiatric PGS (ADHD, autism, bipolar disorder, depression, schizophrenia, anorexia, and insomnia), three personality PGS (extraversion, neuroticism, and openness), three substance use disorders (alcohol dependency, cannabis use disorder, and nicotine dependency), six cognitive PGS (educational attainment, general cognitive performance, executive functioning, non-verbal reasoning, working memory, and reaction time), and BMI. The distributions of these PGS are in Figure S3A, and the correlations between PGS are in Figure S3B.

To detect any potential genetic confounding in our environmental variable (i.e., gene-environment correlates), we first assessed if any PGS were associated with the ADI. We tested for the main effects of PGS on the ADI using weighted linear regression with the formula: *ADI ∼ PGS*. Prior to model input, the ADI was accounted for the 10 genetic principal components (PCs) by linear regression residualization. The *β* estimates with 95% confidence intervals are in Figure 1C. The weighted and ordinary linear regression statistics, heteroscedasticity tests, and Pearson and Spearman correlation coefficients are in Table S1.

Thirteen out of the 20 PGS were significantly associated with the ADI (FDR *p <* 0.05). The strongest association was the educational attainment PGS negatively associated with the ADI (*β* = *−*0.32 [*−*0.34*, −*0.29], adj. *R*^2^ = 0.08), meaning higher environmental disadvantage was associated with lower polygenic propensity for educational attainment. Other cognitive PGS were also negatively associated with the ADI: general cognitive performance (*β* = *−*0.10 [*−*0.13*, −*0.07], adj. *R*^2^ = 0.01) and non-verbal reasoning (*β* = *−*0.03 [*−*0.06, 0], adj. *R*^2^ = 0). The executive functioning PGS was slightly positively associated with the ADI, but not significant after multiple testing correction (*β* = 0.03 [0, 0.06], adj. *R*^2^ = 0, FDR *p* = 0.05). The working memory and reaction time PGS were not associated with the ADI.

The ADI was positively associated with several PGS for psychiatric and substance use disorders, which means that higher environmental disadvantage was associated with higher polygenic risk for these disorders. These significant PGS associations were: ADHD (*β* = 0.17 [0.14, 0.20], adj. *R*^2^ = 0.02), autism (*β* = 0.05 [0.02, 0.08], adj. *R*^2^ = 0), depression (*β* = 0.13 [0.1, 0.16], adj. *R*^2^ = 0.01), insomnia (*β* = 0.11 [0.09, 0.14], adj. *R*^2^ = 0.01), alcohol dependency (*β* = 0.06 [0.04, 0.09], adj. *R*^2^ = 0), cannabis use disorder (*β* = 0.07 [0.04, 0.09], adj. *R*^2^ = 0) and nicotine dependency (*β* = 0.14 [0.12, 0.17], adj. *R*^2^ = 0.01). Bipolar disorder and schizophrenia were not significantly associated with the ADI. Interestingly, the anorexia PGS was negatively associated with the ADI (*β* = *−*0.04 [*−*0.06*, −*0.01], adj. *R*^2^ = 0). Of the three personality PGS, openness and extraversion were not associated with the ADI, while neuroticism was positively associated (*β* = 0.10 [0.07, 0.12], adj. *R*^2^ = 0.01). Additionally, the BMI PGS was positively associated with the ADI (*β* = 0.17 [0.15, 0.20], adj. *R*^2^ = 0.02).

The PGS associations with the ADI were similar when calculated separately for ABCD and SPARK (Figure S4).

#### 3.1.4 SNP-based heritability of the Area Deprivation Index

Because the ADI was associated with many PGS, we calculated the SNP-based heritability (*h*^2^) to determine the total variance explained by the common SNPs in our dataset (Figure 1D). We used GCTA-REML to calculate *h*^2^ with the 10 genetic PCs as covariates. The ADI had significant SNP heritability: *h*^2^ = 0.33 [0.24, 0.42]*, p <* 0.05. This estimate was relatively similar when calculated separately for each cohort: ABCD *h*^2^ = 0.28 [0.11, 0.45]*, p <* 0.05 and SPARK *h*^2^ = 0.20 [0.02, 0.39], *p <* 0.05. Because our sample is children, we reasoned that the PGS associations with the ADI are capturing genetic nurture [54]. In order to account for these genetic nurture effects in subsequent analyses, we residualized the 20 PGS from the ADI using linear regression residualization. This reduced the heritability, but the ADI still had significant heritability: *h*^2^ = 0.15 [0.06, 0.24], *p <* 0.05. This indicates that the *h*^2^ of the ADI is only partially accounted for by linear combinations of the 20 PGS and that there is further genetic signal remaining after the PGS are residualized from the ADI.

### 3.2 Independent effects of the Area Deprivation Index and polygenic scores on behavioral problems

#### 3.2.1 Measures of behavioral problems

The estimates of behavioral problems were the five syndromic subscales from the Child Behavior Checklist (CBCL). The CBCL is a parent-report questionnaire of 118 items that covers a wide range of behavioral and emotional problems that is appropriate for children aged 6 to 17 years. Specific items are summed to form five quantitative syndromic subscales: attention problems, social problems, thought problems, internalizing problems (anxious, withdrawn depressed, and somatic complaints), and externalizing problems (aggressive and rule-breaking behavior). The distributions of these five CBCL subscales in our sample are in Figure S1C. The scores were higher in SPARK than in ABCD. The final scores that were used as phenotypes in all subsequent analyses were accounted for age in months by quasi-Poisson regression residualization and then Z-scaled (Figure S1D).

#### 3.2.2 Independent effects of the Area Deprivation Index and polygenic scores on behavioral problems

Behavioral problems in children are influenced by both environmental and genetic factors, but head-to-head comparisons of environmental adversity versus several PGS have not yet been investigated. Therefore, we first tested for the independent effects of the ADI and 20 PGS on the CBCL scores (*CBCL ∼ ADI* and *CBCL ∼ PGS*). Figure 2 shows the adjusted *R*^2^ from these weighted linear regression models, and Tables S2 and S3 have the ordinary regression statistics and correlation statistics.

**Figure 2.**
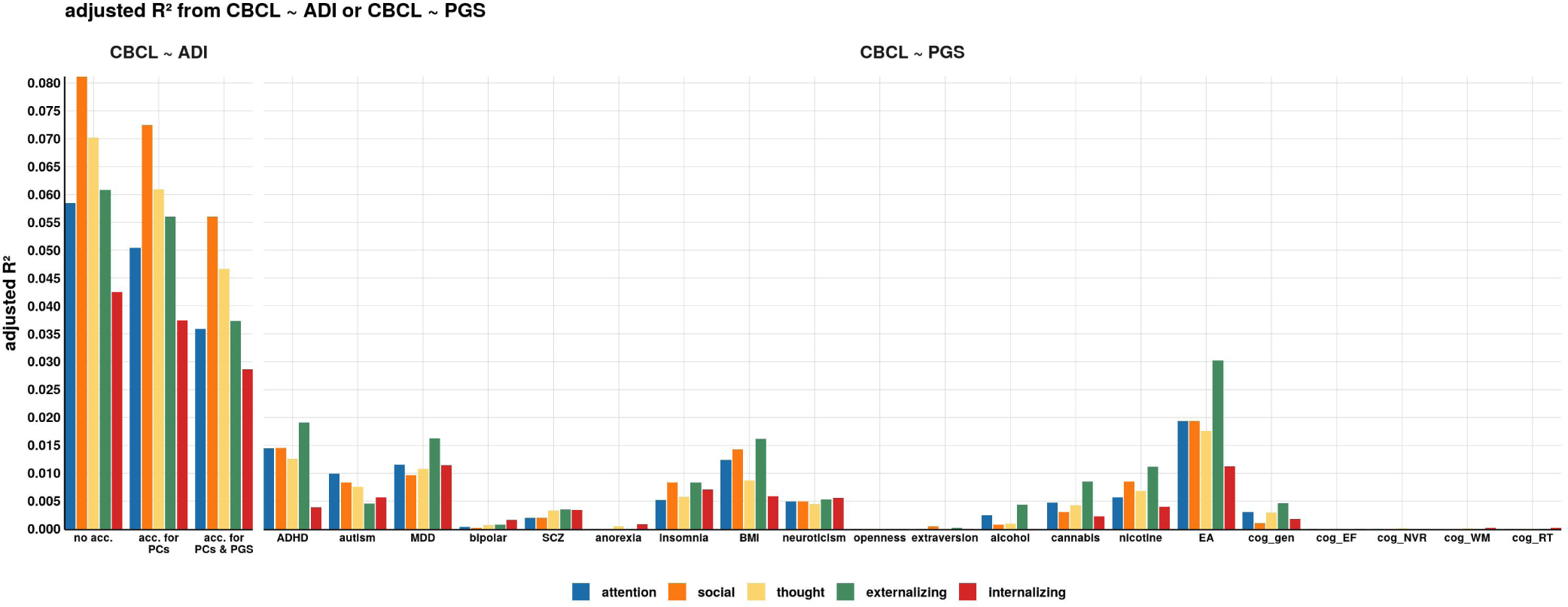
Comparison of variance explained by the independent effects of the Area Deprivation Index versus polygenic scores. Adjusted *R*^2^ from weighted least squares linear regression models testing the independent effects of the Area Deprivation Index (ADI) and the 20 polygenic scores (PGS) on the Child Behavior Checklist (CBCL) subscale scores (*CBCL ∼ ADI* and *CBCL ∼ PGS*). The full model statistics are in Table S2 and Table S3. The models calculated separately for ABCD and SPARK are in Figure S5. PGS: **MDD** = major depressive disorder, **SCZ** = schizophrenia, **alcohol** = alcohol dependency, **cannabis** = cannabis use disorder, **nicotine** = nicotine dependency, **EA** = educational attainment, **cog gen** = general cognitive performance, **cog EF** = executive functioning, **cog NVR** = non-verbal reasoning, **cog WM** = working memory, **cog RT** = reaction time.

The ADI was positively associated with the five CBCL scores, meaning that higher environmental adversity was associated with more behavioral problems. The variance explained by the ADI was highest when the ADI was not accounted for the 10 genetic PCs nor the 20 PGS, with the adjusted *R*^2^ ranging from 0.04 for internalizing problems to 0.08 for social problems. Accounting for the 10 genetic PCs slightly reduced the adjusted *R*^2^ to 0.07 for social problems (the adjusted *R*^2^ for internalizing problems stayed at 0.04). Accounting for the 10 genetic PCs and the 20 PGS further reduced the adjusted *R*^2^ to 0.03 for internalizing problems and 0.06 for social problems.

The variance explained by the ADI was higher than the variance explained by any of the 20 PGS. The highest adjusted *R*^2^ was the educational attainment PGS negatively associated with externalizing problems (adj. *R*^2^ = 0.03, FDR *p <* 0.05, *β* = *−*0.18 [*−*0.20*, −*0.16]). The variance explained by the ADHD PGS for externalizing problems was similar (adj. *R*^2^ = 0.02, FDR *p <* 0.05, *β* = 0.14 [0.12, 0.17]). Internalizing problems was CBCL subscale least explained by any PGS. The strongest associations were the educational attainment PGS (adj. *R*^2^ = 0.01, FDR *p <* 0.05, *β* = *−*0.11 [*−*0.13*, −*0.09]) and the depression PGS (adj. *R*^2^ = 0.01, FDR *p <* 0.05, *β* = 0.11 [0.09, 0.13]).

The models calculated separately for ABCD and SPARK are in Figure S5.

### 3.3 Interaction effects (GxE) of the Area Deprivation Index and polygenic scores

Next, we investigate whether the ADI and PGS have additive and interaction effects on CBCL scores by including both independent variables in the models plus the interaction term using weighted linear regression: *CBCL ∼ ADI* + *PGS* + *ADI × PGS*. We tested for interaction before the ADI was accounted for the PGS (Figure S6 and after the ADI was accounted for the PGS. The main results report the effects after the ADI was accounted for the PGS. Figure 3 shows the *β* estimates for each of the three terms (*ADI*, *PGS*, and *ADI × PGS*). Also see Table S4 for all model statistics.

**Figure 3.**
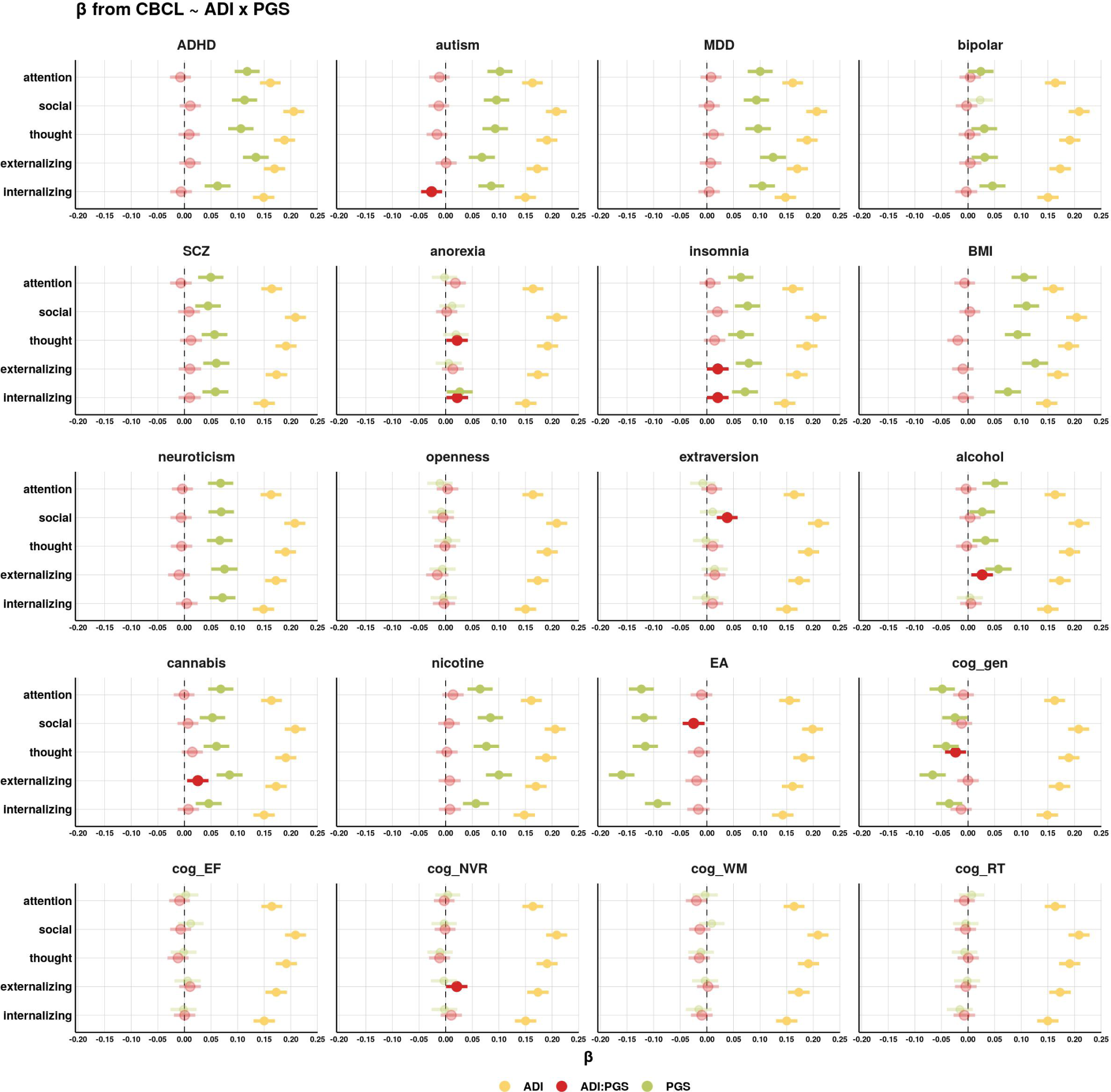
Interaction effects of the Area Deprivation Index and polygenic scores on behavioral problems. The additive effects of the Area Deprivation Index (ADI), polygenic scores (PGS), and the ADI-by-PGS interaction effect were modeled using weighted linear regression: *CBCL ∼ ADI* + *PGS* + *ADI × PGS*. Prior to model input, the ADI was accounted for the 10 genetic principal components (PCs) and 20 PGS by linear regression residualization. The *β* estimates are with the 95% confidence intervals. *β* estimates that are nominally significant (unadjusted *p <* 0.05) have a solid fill color. We also ran models with the ADI not residualized for the 20 PGS prior to model input (Figure S6). Additionally, the results calculated separately for ABCD and SPARK are in Figure S7. PGS: **MDD** = major depressive disorder, **SCZ** = schizophrenia, **alcohol** = alcohol dependency, **cannabis** = cannabis use disorder, **nicotine** = nicotine dependency, **EA** = educational attainment, **cog gen** = general cognitive performance, **cog EF** = executive functioning, **cog NVR** = non-verbal reasoning, **cog WM** = working memory, **cog RT** = reaction time.

Many PGS contributed additively with the ADI, with some of the PGS effects similar in magnitude to the ADI effect. For attention problems the ADI effect was: *β* = 0.16 [0.14, 0.18], FDR *p <* 0.05 and the ADHD PGS effect was: *β* = 0.12 [0.09, 0.14], FDR *p <* 0.05. For internalizing problems the ADI effect was *β* = 0.15 [0.13, 0.17], FDR *p <* 0.05 and the depression PGS effect was *β* = 0.10 [0.08, 0.13], FDR *p <* 0.05.

Other psychiatric PGS had nominally significant ADI-by-PGS interactions (GxE) in addition to the main effects of the PGS. The insomnia PGS had two significant ADI-by-PGS interaction effects in which the interaction effect went the same direction as the main effect of the PGS, meaning increasing ADI amplified the PGS effect. Specifically, for externalizing problems, the insomnia PGS interaction effect was positive (*β* = 0.02 [0, 0.04]*, p* = 0.05, FDR *p* = 0.4), as was the PGS main effect (*β* = 0.08 [0.05, 0.1], FDR *p <* 0.05). The anorexia PGS also had two nominally significant interactions for thought problems and internalizing problems. For thought problems, the anorexia PGS interaction effect was positive (*β* = 0.02 [0, 0.04]*, p* = 0.04, FDR *p* = 0.40) and the main effect was positive but not significant (*β* = 0.02 [0, 0.04]*, p* = 0.11). For internalizing problems, the anorexia PGS interaction effect was positive (*β* = 0.02 [0, 0.04]*, p* = 0.04, FDR *p* = 0.40), and PGS main effect was also positive (*β* = 0.03 [0, 0.05]*, p* = 0.03). Interestingly, for internalizing problems, the autism PGS interaction effect was negative (*β* = *−*0.03 [*−*0.05*, −*0.01]*, p* = 0.01, FDR *p* = 0.33), but the PGS main effect was positive (*β* = 0.09 [0.06, 0.11], FDR *p <* 0.05). This means that an increasing ADI *suppressed* the autism PGS effect. No other psychiatric PGS had significant interaction effects.

The personality and substance use disorder PGS had interaction trends with more problem-type specificity. The extraversion PGS had a strong positive interaction effect for social problems (*β* = 0.04 [0.02, 0.06], FDR *p <* 0.05), with the PGS main effect not significant (*β* = 0.01 [*−*0.01, 0.03]). For the openness PGS, the interaction effects were all *β* = 0 except for externalizing problems: *β* = *−*0.02 [*−*0.04, 0.01]*, p* = 0.14. The neuroticism PGS did not have any interaction effects. The alcohol and cannabis dependency PGS had significant interactions for externalizing problems only. These interaction effects were positive for alcohol dependency (*β* = 0.03 [0.01, 0.05]*, p* = 0.01, FDR *p* = 0.33) and cannabis dependency (*β* = 0.03 [0.01, 0.05]*, p* = 0.01, FDR *p* = 0.33).

The educational attainment PGS had significant (FDR *p <* 0.05) negative main effects on all five CBCL subscales, with the effects ranging from *β* = *−*0.09 [*−*0.12*, −*0.07] for internalizing problems to *β* = *−*0.16 [*−*0.18*, −*0.14] for externalizing problems. The educational attainment PGS interaction effect was significant for social problems and went in the same direction as the PGS main effect *β* = *−*0.02 [*−*0.05, 0]*, p* = 0.02, FDR *p* = 0.33. The educational attainment PGS interaction effects on the other four CBCL subscales were in the same direction but not nominally significant. For thought problems, the general cognitive performance PGS main effect was negative (*β −* 0.04 [*−*0.07*, −*0.02], FDR *p <* 0.05) as was the interaction effect (*β* = *−*0.02 [*−*0.04, 0]*, p* = 0.02, FDR *p* = 0.33).

The results calculated separately for ABCD and SPARK are in Figure S7.

### 3.4 Average marginal effects of polygenic scores across the range of the Area Deprivation Index

Penetrance is the extent to which the PGS is expressed in a given environment, or, in other words, how the PGS effect changes across the environment. To further interrogate the GxE effects and tailor the analysis towards a more traditional genetic framework, we calculated the average marginal effects (AME) of PGS across the ADI, which effectively captures the PGS penetrance. AME calculates the average effect of one independent variable (the PGS) at different fixed values of the other independent variable (the ADI). One advantage of using the ADI is that we know that we have captured the full range of the environmental variable. We calculated the AME of the PGS at every 0.5 standard deviation of the ADI using weighted least squares linear regression. Prior to model input, the ADI was accounted for the 10 genetic PCs and the 20 PGS by linear regression residualization and then Z-scaled (i.e., mean (*µ*) of 0 and a standard deviation (*σ*) of 1). This ADI had a minimum of -2.45 and a maximum of 3.60. Figure 4 shows the AME of the PGS across the ADI, with complete statistics in Table S5.

**Figure 4.**
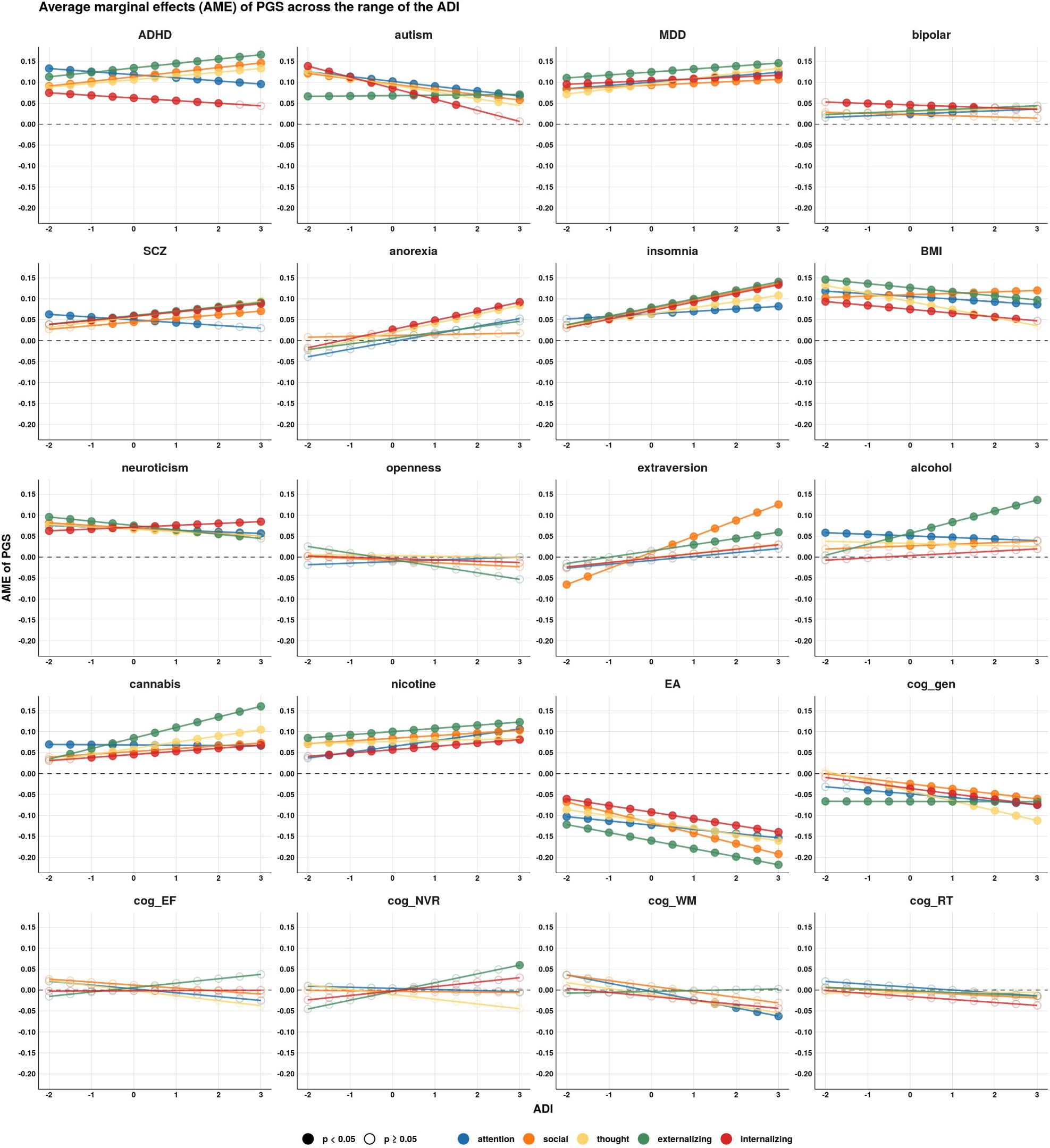
Average marginal effects of polygenic scores across the range of the Area Deprivation Index. Average marginal effects (AME) of the 20 polygenic scores (PGS) were calculated along the range of the Area Deprivation Index (ADI) at every 0.5 value of the ADI using weighted least squares linear regression. Prior to model input, the ADI was accounted for the 10 genetic principal components (PCs) and the 20 PGS by linear regression residualization. AME of PGS that were nominally significant (unadjusted *p <* 0.05) have a solid fill color. The AME calculated with the ADI not residualized for the 20 PGS are in Figure S8. The AME calculated separately for ABCD and SPARK are in Figure S9. PGS: **MDD** = major depressive disorder, **SCZ** = schizophrenia, **alcohol** = alcohol dependency, **cannabis** = cannabis use disorder, **nicotine** = nicotine dependency, **EA** = educational attainment, **cog gen** = general cognitive performance, **cog EF** = executive functioning, **cog NVR** = non-verbal reasoning, **cog WM** = working memory, **cog RT** = reaction time.

The autism PGS had a significant interaction effect on internalizing problems, with the interaction effect going the opposite direction of the main effect. The AME of the autism PGS in an advantaged environment (ADI = -2) was *β* = 0.14 [0.09, 0.19], but in a disadvantaged environment (ADI = 3) the autism PGS AME was *β* = 0.01 [*−*0.05, 0.06]. In other words, the autism PGS was associated with more internalizing problems *only* in an advantaged environment. In a disadvantaged environment, the autism PGS was not associated with internalizing problems. This means the penetrance of the autism PGS on internalizing problems decreased as the ADI increased. The other psychiatric PGS with significant interaction effects had the opposite effect, that is, the AME of the PGS was greatest in a disadvantaged environment. The anorexia PGS AME on thought problems in an advantaged environment was *β* = *−*0.02 [*−*0.08, 0.03], but in a disadvantaged environment the AME increased to *β* = 0.08 [0.03, 0.14]. The insomnia PGS AME on externalizing problems in an advantaged environment was *β* = 0.04 [*−*0.02, 0.09] but in a disadvantaged environment the AME was *β* = 0.14 [0.08, 0.20].

The AME of the alcohol and cannabis dependency PGS on externalizing problems increased as the ADI increased. In an advantaged environment, the AME of the alcohol dependency PGS was *β* = 0 [*−*0.05, 0.06], but in a disadvantaged environment the AME was *β* = 0.14 [0.08, 0.19]. The effect was similar for the cannabis dependency PGS; in an advantaged environment, the AME was *β* = 0.03 [*−*0.02, 0.09], but in a disadvantaged environment the AME was *β* = 0.16 [0.10, 0.22]. Interestingly, the AME of the extraversion PGS on social problems changed sign across the ADI. In an advantaged environment, the AME was *β* = *−*0.07 [*−*0.12*, −*0.01], but in a disadvantaged environment the AME was *β* = 0.13 [0.07, 0.18].

The educational attainment PGS effect increased as the ADI increased, most prominently for social problems. In an advantaged environment, the AME was *β* = *−*0.07 [*−*0.12*, −*0.10], but in a disadvantaged environment the AME was *β* = *−*0.19 [*−*0.25*, −*0.13]. Similarly for externalizing problems, in an advantaged environment the AME was *β* = *−*0.12 [*−*0.18*, −*0.07], but in a disadvantaged environment the AME was *β* = *−*0.22 [*−*0.28*, −*0.16]. The AME of the general cognitive performance PGS on thought problems in an advantaged environment was *β* = 0.01 [*−*0.05, 0.06], but in a disadvantaged environment the AME was *β* = *−*0.11 [*−*0.17*, −*0.06]. Surprisingly, for the non-verbal reasoning PGS effect on externalizing problems, the AME effect in an advantaged environment was *β* = *−*0.05 [*−*0.10, 0.01], but in a disadvantaged environment the AME was positive *β* = 0.06 [0, 0.12]. The AME were also calculated when the ADI was not residualized for the 20 PGS (Figure S8). The AME were also calculated separately for ABCD and SPARK (Figure S9).

## 4 Discussion

In this GxE study, we investigated how the Area Deprivation Index (ADI) moderates the effects of 20 polygenic scores (PGS) on five estimates of behavioral problems in *N* = 7, 191 children in the United States. The combination of the ABCD and SPARK cohorts allowed us to have a greater range in the various behavioral problems due to the enrichment of neurodevelopmental and psychiatric conditions in the SPARK autism cohort. Additionally, our study was strengthened by using continuous, dimensional estimates of behavior [19, 55], as well as continuous estimates of environment and genetic variables.

We found the ADI to have significant confounding genetic signal (i.e., gene-environment correlates). Thirteen out of the 20 PGS were significantly associated with the ADI (Figure 1C). Psychiatric and substance use disorder PGS were positively associated with ADI, meaning that an increased polygenic risk for these disorders was associated with greater environmental disadvantage. The educational attainment PGS had the strongest negative association, with lower educational attainment PGS associated with greater environmental disadvantage. Other cognitive PGS were also negatively associated with the ADI but to a lesser extent. The neighborhood locations of our participants are reflective of their parents’ histories/decisions (not the participants’, as they are children). Therefore, the PGS associations with the ADI may be capturing the effects of genetic nurture [54]. Previous work has found genetic nurture effects to be especially influential on educational outcomes [56]. Because we reasoned that the PGS associations with the ADI are capturing genetic nurture, we accounted for these effects in subsequent analyses by residualizing the 20 PGS from the ADI. This reduced the SNP heritability of the ADI, but it still had significant heritability after residualizing for the PGS (Figure 1D). It is not surprising that the genetic signal contained by the ADI is not completely explained by linear combinations of the 20 PGS. The significant genetic signal we identified in the ADI underscores the importance of genetic variables in sociological and public policy research [57] and should warrant caution in interpreting the ADI as a purely environmental variable.

In head-to-head comparisons of the effects of the ADI versus PGS on behavioral problems, we found the ADI to out-perform all 20 PGS (Figure 3). However, accounting the ADI for the 20 PGS reduced the adjusted *R*^2^. Out of all the PGS, the educational attainment, ADHD, and depression PGS were most associated with CBCL scores. When additively modeling both the ADI and the PGS with an interaction term (*CBCL ∼ ADI* + *PGS* + *ADI × PGS*), we found the ADI and many PGS to have independent, additive effects and a few to have ADI-by-PGS interaction effects (Figure 3). Even when using the ADI that was not accounted for the 20 PGS, many PGS had effects that were independent of the ADI effect (Figure S6). These results demonstrate that although the ADI is more predictive than PGS, the PGS contribute additional effects that are not captured by the ADI. This is in line with previous work of twins in ABCD (*N* = 772 pairs) which found that many traits, including the CBCL subscales, had independent genetic and environmental effects [58].

Because the ADI is continuous data, we were able to confirm that the full range of this environmental variable was represented in our sample, which is crucial for GxE studies [59]. We identified several ADI-by-PGS interaction effects (GxE). Almost all of these interaction effects were in the same direction as the PGS main effect, meaning the ADI amplified the PGS main effect (i.e., the PGS penetrance increased as environmental disadvantage increased). For example, we found that the positive average marginal effect (AME) of substance use disorder PGS on externalizing problems increased as the environmental disadvantage increased (Figure 4). This reflects the inherited sensitivity mechanism (also known as the diathesis-stress mechanism) in which the environment triggers a genetic susceptibility and therefore the genetic penetrance, or heritability, is highest in a stressful environment [59]. Interestingly, our results are in contrast to a recent study of European twins that showed the heritability of childhood psychotic experiences decreased with increasing environmental exposures [60]. However, this study used different methodology and only looked at psychotic phenotypes, which we did not use (the CBCL thought problems subscale is the closest analog to reported psychotic experiences [61]). The only GxE effect we found that showed decreased PGS penetrance as the ADI increased was the autism PGS effect on internalizing problems. This suggests that the autism PGS is predictive of internalizing problems *only* in an advantaged environment.

### 4.1 Limitations

This study has several limitations. First, the GWAS sample populations that we used to calculate the PGS are from samples of European ancestry with known participation biases like the UK Biobank, and these participation biases have genetic effects [62]. Because the GWAS samples only included European genetic ancestry, we regrettably were only able to include participants from ABCD and SPARK with European genetic ancestry. Future work needs to perform GWAS in diverse populations so that all participants can be included for PGS analysis. Second, our linear models violated the homoscedasticity assumption (i.e., the CBCL variance was not constant across the ADI). This violation is not uncommon in GxE models. New conceptual frameworks are being developed to articulate the distinction between a significant GxE effect due to the environmental variable amplifying/suppressing the PGS effect or the environmental variable amplifying the total variation in the phenotype [63]. We addressed the heteroscedasticity problem with weighted linear regression in which each percentile of the ADI contributed equally to the models. This did reduce the heteroscedasticity but did not resolve it (Table S2). Additionally, we also ran non-parametric Spearman correlations (which do not require homoscedasticity) and did not identify any substantial deviations (Table S1, Table S2, and Table S3). Third, while combining ABCD and SPARK was advantageous in regards to expanding our phenotypic and genetic variability, it also introduced the limitation that these are separate cohorts with different enrollment criteria. Additionally, SPARK is an autism cohort, so it is likely that SPARK is enriched for rare deleterious variants and copy number variants [64, 65], especially compared to ABCD.

## Data Availability

The SPARK genetic data can be obtained at SFARI Base: https://base.sfari.org. Data used in the preparation of this article were obtained from the Adolescent Brain Cognitive Development (ABCD) Study (https://abcdstudy.org), held in the NIMH Data Archive (NDA).

## Acknowledgments

We are grateful to all of the participants and families in ABCD and SPARK, the SPARK clinical sites, and SPARK staff. We appreciate obtaining access to genetic and phenotypic data for SPARK data on SFARI Base.

## Data availability

The SPARK genetic data can be obtained at SFARI Base: https://base.sfari.org Data used in the preparation of this article were obtained from the Adolescent Brain Cognitive Development (ABCD) Study (https://abcdstudy.org), held in the NIMH Data Archive (NDA). This is a multisite, longitudinal study designed to recruit more than 10,000 children age 9-10 and follow them over 10 years into early adulthood. The ABCD Study is supported by the National Institutes of Health and additional federal partners under award numbers U01DA041048, U01DA050989, U01DA051016, U01DA041022, U01DA051018, U01DA051037, U01DA050987, U01DA041174, U01DA041106, U01DA041117, U01DA041028, U01DA041134, U01DA050988, U01DA051039, U01DA041156, U01DA041025, U01DA041120, U01DA051038, U01DA041148, U01DA041093, U01DA041089, U24DA041123, U24DA041147. A full list of supporters is available at https://abcdstudy.org/federal-partners.html. A listing of participating sites and a complete listing of the study investigators can be found at https://abcdstudy.org/consortium_members/. ABCD consortium investigators designed and implemented the study and/or provided data but did not necessarily participate in the analysis or writing of this report. This manuscript reflects the views of the authors and may not reflect the opinions or views of the NIH or ABCD consortium investigators. The ABCD data repository grows and changes over time. The ABCD data used in this report came from [NIMH Data Archive Digital Object Identifier 10.15154/1523041]. DOIs can be found at https://nda.nih.gov/abcd. Additional support for this work was made possible from NIEHS R01-ES032295 and R01-ES031074.

## Author contributions

TRT contributed to the study design, generating the polygenic scores, processing the data, performing the statistical analyses, and writing the manuscript. LCG contributed to processing the data and writing the manuscript. JJM contributed to study design and writing the manuscript.

## Supplementary information

**Figure S1.**
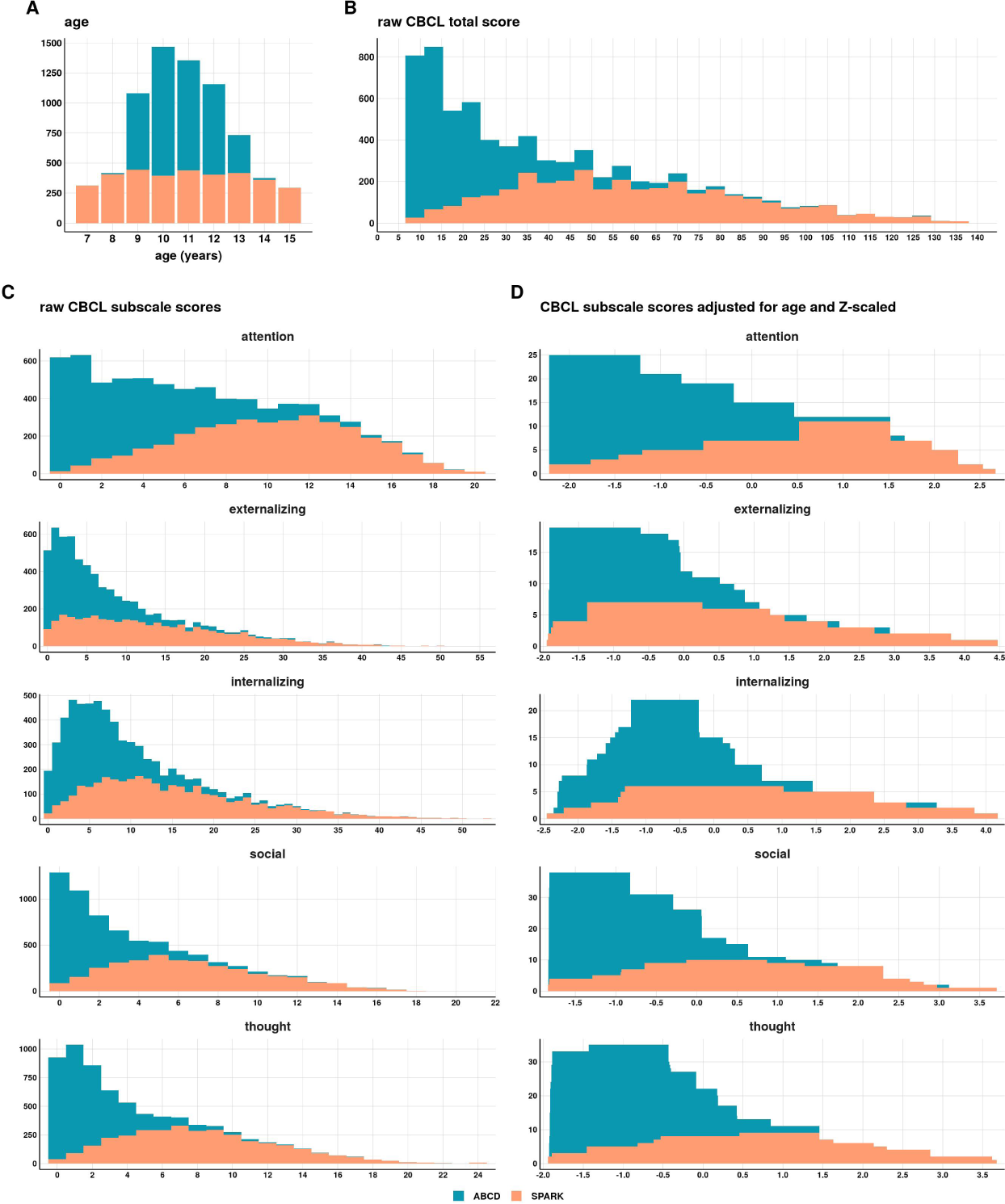
Distributions of ages and the Child Behavior Checklist subscale scores. **(A)** Distribution of age in our sample (*N* = 7, 191). **(B)** Distribution of the Child Behavior Checklist (CBCL) total score. We removed participants that were extreme outliers (above the 99.5^th^ percentile in our sample, which was a score of 7). As per recommendations of [19], we were strict with our inclusion criteria for low total scores. Therefore, we removed participants that were below the 20^th^ percentile (a score of 7). The behavioral problem phenotypes were the five syndromic subscales from the CBCL: attention problems, social problems, thought problems, externalizing problems, and internalizing problems. **(C)** Distributions of the raw CBCL subscale scores. **(D)** Distributions of the CBCL subscale scores after accounting for age by quasi-Poisson regression residualization and then Zscaling (*µ* = 0 *σ* = 1). These age-adjusted scores were used as the phenotypes in all subsequent analyses.

**Figure S2.**
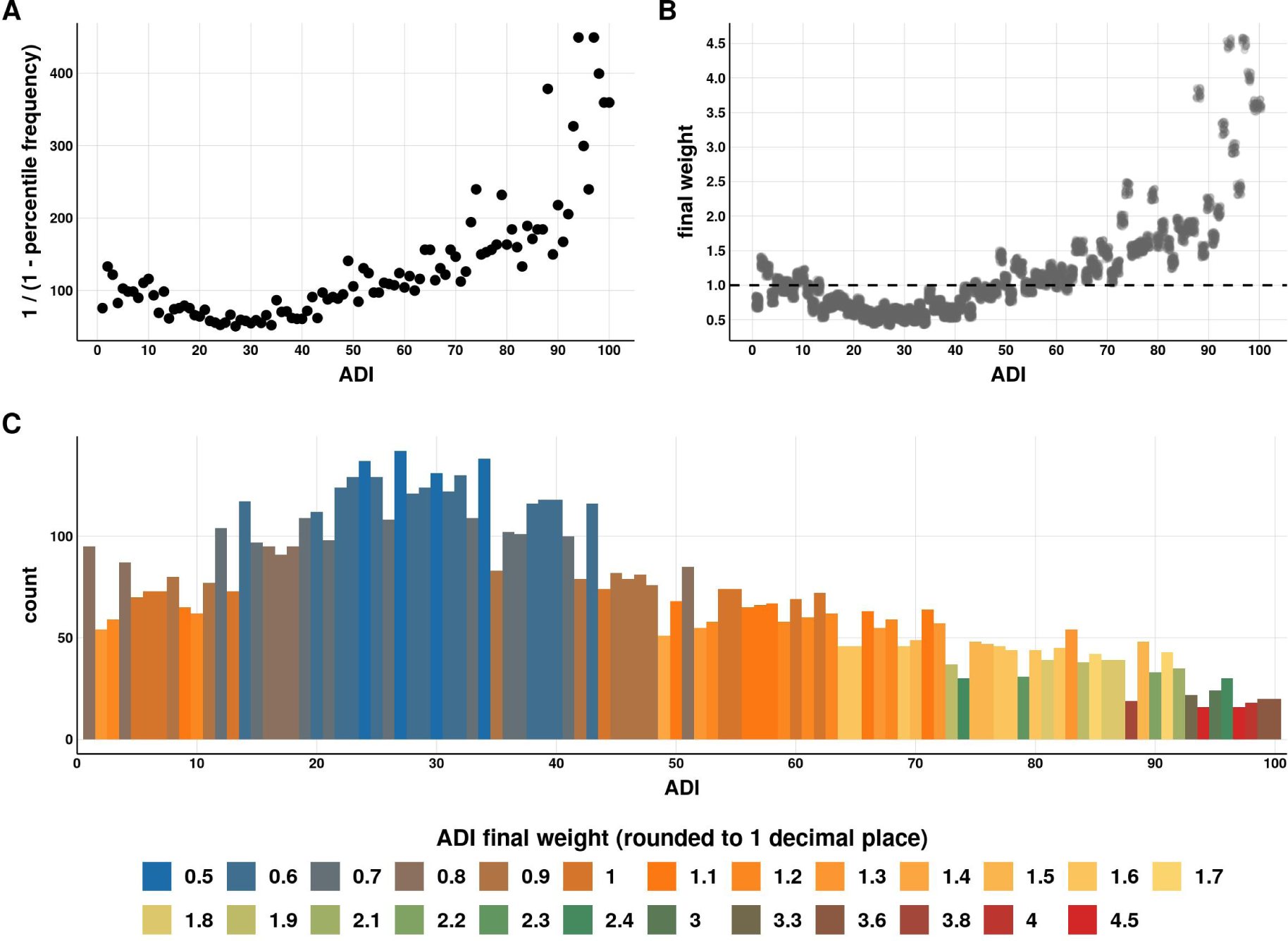
Generation of weights from the Area Deprivation Index distribution. The Area Deprivation Index (ADI) is continuous percentile data, so the distribution should be uniform (i.e., equal N per ADI percentile). Our sample was not uniform across the ADI. We used our sample distribution to generate weights so that each ADI percentile contributed equally to subsequent linear regression models. **(A)** For each percentile, we first took the inverse of one minus the frequency. **(B)** These values were then assigned to each sample based on their ADI and then rescaled so that the mean final weight was 1 and the sum of weights was equal to the number of samples (*N* = 7, 191). **(C)** The weights ranged from 0.5 to 4.5. Samples with the highest weights are filled red and are in higher ADI percentiles (ADI *>* 95) whereas the samples with lowest weights are filled blue and are in ADI percentiles ranging from approximately 20 to 40.

**Figure S3.**
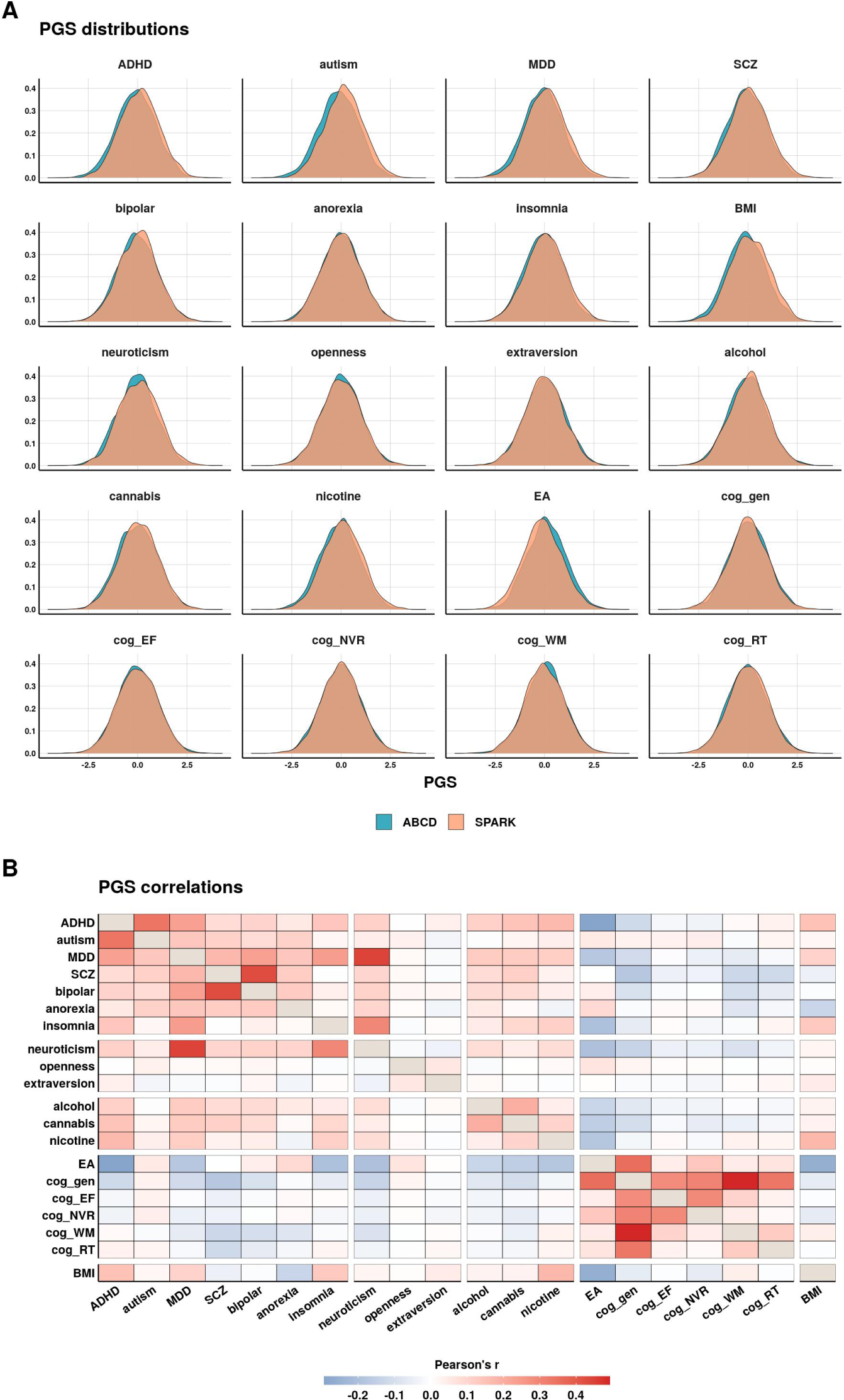
Polygenic score distributions and correlations. **(A)** Distributions of the 20 polygenic scores (PGS) after accounting for the major axes of genetic variation by residualizing the PGS for the 10 genetic principal components and then Z-scaling (*µ* = 0*, σ* = 1). **(B)** Correlations between the 20 PGS. The fill color is Pearson’s *r*. PGS: **MDD** = major depressive disorder, **SCZ** = schizophrenia, **alcohol** = alcohol dependency, **cannabis** = cannabis use disorder, **nicotine** = nicotine dependency, **EA** = educational attainment, **cog gen** = general cognitive performance, **cog EF** = executive functioning, **cog NVR** = non-verbal reasoning, **cog WM** = working memory, **cog RT** = reaction time.

**Figure S4.**
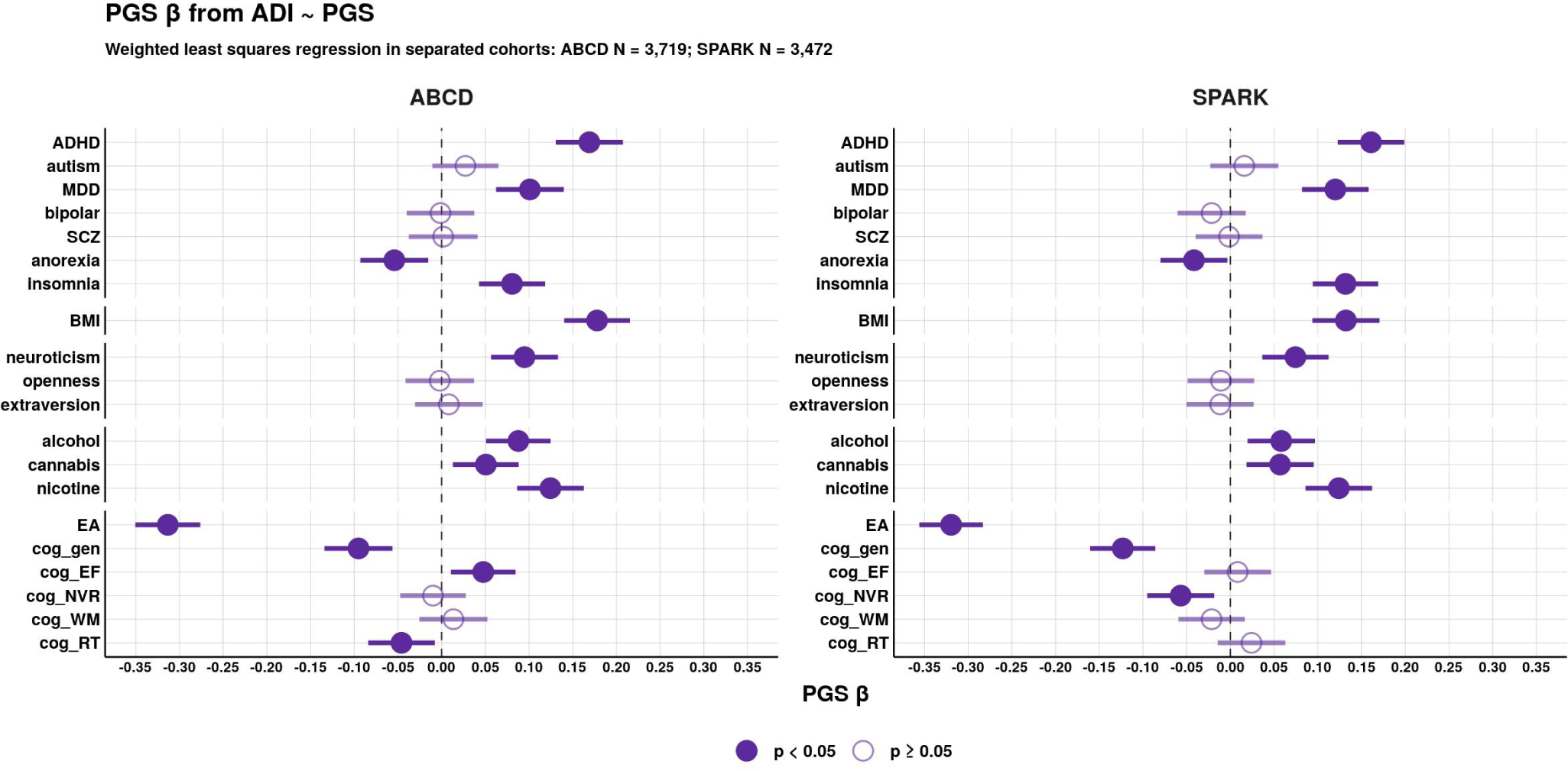
Polygenic score associations with the Area Deprivation Index calculated separately for ABCD and SPARK. Associations of 20 polygenic scores (PGS) with the Area Deprivation Index (ADI). The PGS *β* estimates are with the 95% confidence intervals from the weighted linear regression model: *ADI ∼ PGS*. Models were calculated separately between the two cohorts: ABCD *N* = 3, 719 and SPARK *N* = 3, 472, with the weights re-calculated for each cohort. PGS: **MDD** = major depressive disorder, **SCZ** = schizophrenia, **alcohol** = alcohol dependency, **cannabis** = cannabis use disorder, **nicotine** = nicotine dependency, **EA** = educational attainment, **cog gen** = general cognitive performance, **cog EF** = executive functioning, **cog NVR** = non-verbal reasoning, **cog WM** = working memory, **cog RT** = reaction time.

**Figure S5.**
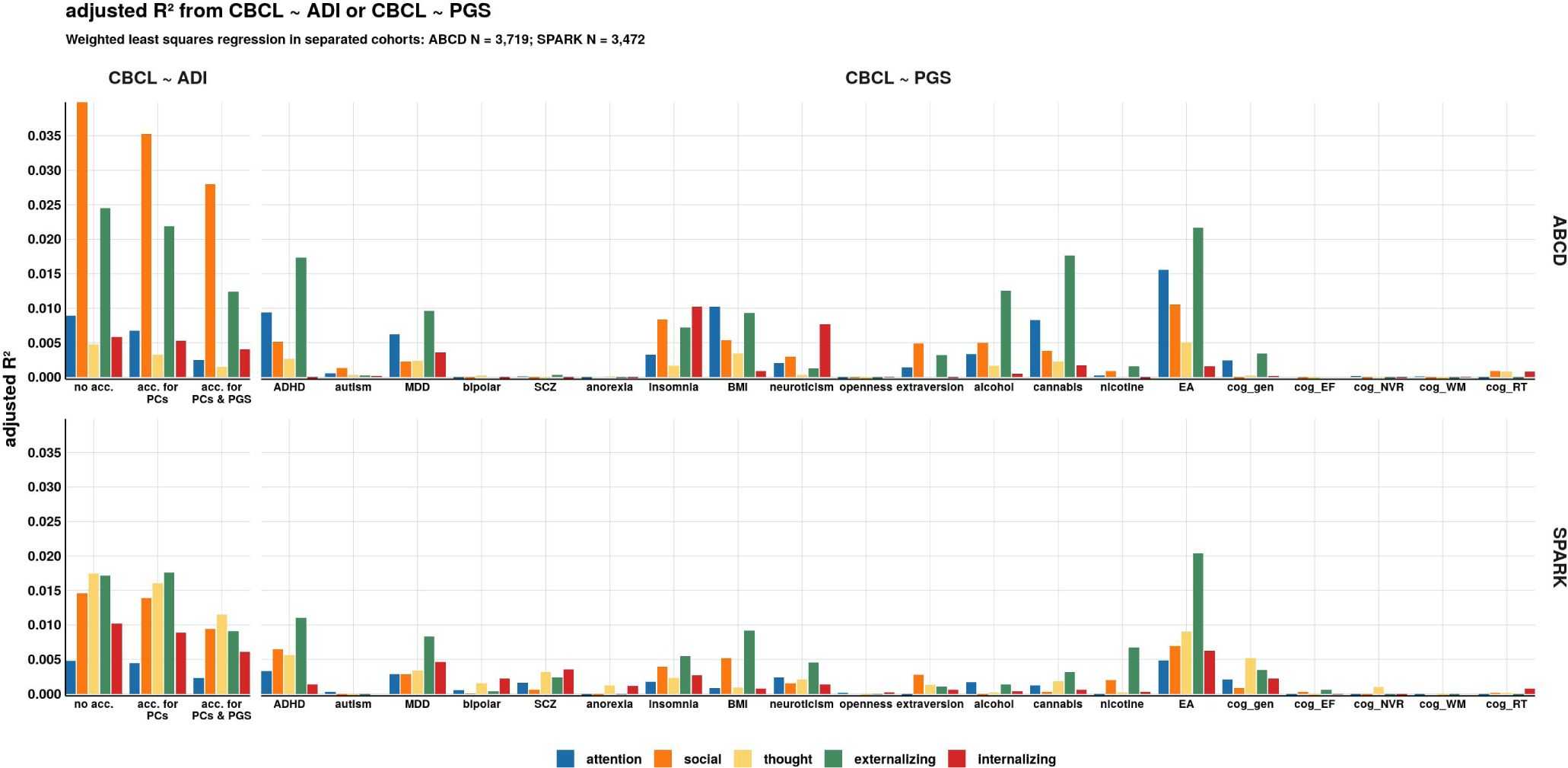
Comparison of the variance explained by the Area Deprivation Index versus the polygenic scores calculated separately for ABCD and SPARK. Adjusted *R*^2^ from weighted least squares linear regression models testing the independent effects of the Area Deprivation Index (ADI) and the 20 polygenic scores (PGS) on the Child Behavior Checklist (CBCL) subscale scores (*CBCL ∼ ADI* and *CBCL ∼ PGS*). Models were calculated separately between the two cohorts: ABCD *N* = 3, 719 and SPARK *N* = 3, 472, with the weights re-calculated for each cohort. PGS: **MDD** = major depressive disorder, **SCZ** = schizophrenia, **alcohol** = alcohol dependency, **cannabis** = cannabis use disorder, **nicotine** = nicotine dependency, **EA** = educational attainment, **cog gen** = general cognitive performance, **cog EF** = executive functioning, **cog NVR** = non-verbal reasoning, **cog WM** = working memory, **cog RT** = reaction time.

**Figure S6.**
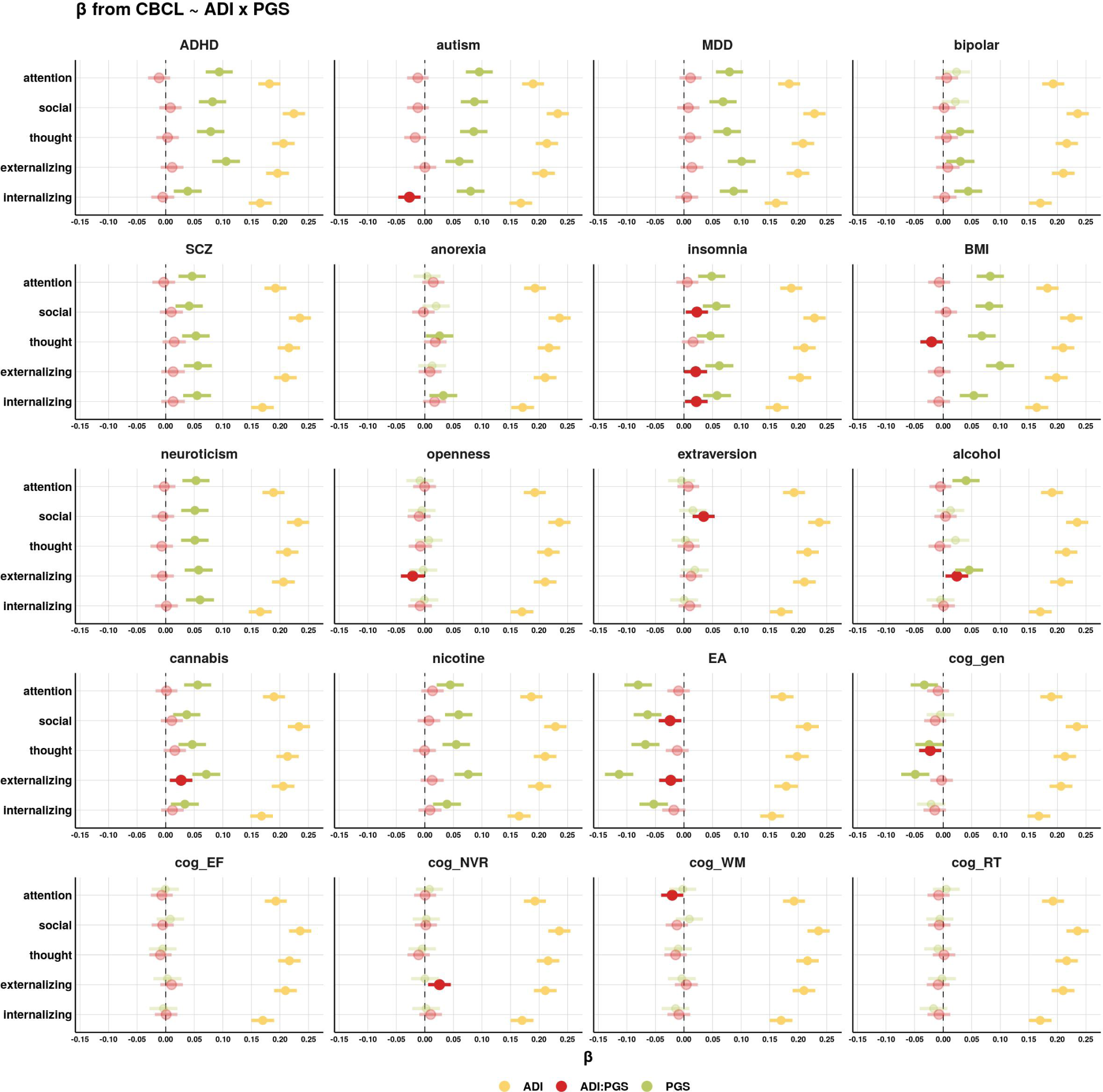
Interaction effects of the Area Deprivation Index and polygenic scores on behavioral problems. The Area Deprivation Index was not accounted for the 20 polygenic scores. The additive effects of the Area Deprivation Index (ADI), polygenic scores (PGS), and the ADI-by-PGS interaction effect were modeled using weighted linear regression: *CBCL ∼ ADI* + *PGS* + *ADI × PGS*. The ADI was not accounted for the 20 PGS, but was residualized for the 10 genetic principal components (PCs) and Z-scaled (*µ* = 0*, σ* = 1). The *β* estimates are with the 95% confidence intervals. *β* estimates that are nominally significant (unadjusted *p <* 0.05) have a solid fill color. PGS: **MDD** = major depressive disorder, **SCZ** = schizophrenia, **alcohol** = alcohol dependency, **cannabis** = cannabis use disorder, **nicotine** = nicotine dependency, **EA** = educational attainment, **cog gen** = general cognitive performance, **cog EF** = executive functioning, **cog NVR** = non-verbal reasoning, **cog WM** = working memory, **cog RT** = reaction time.

**Figure S7.**
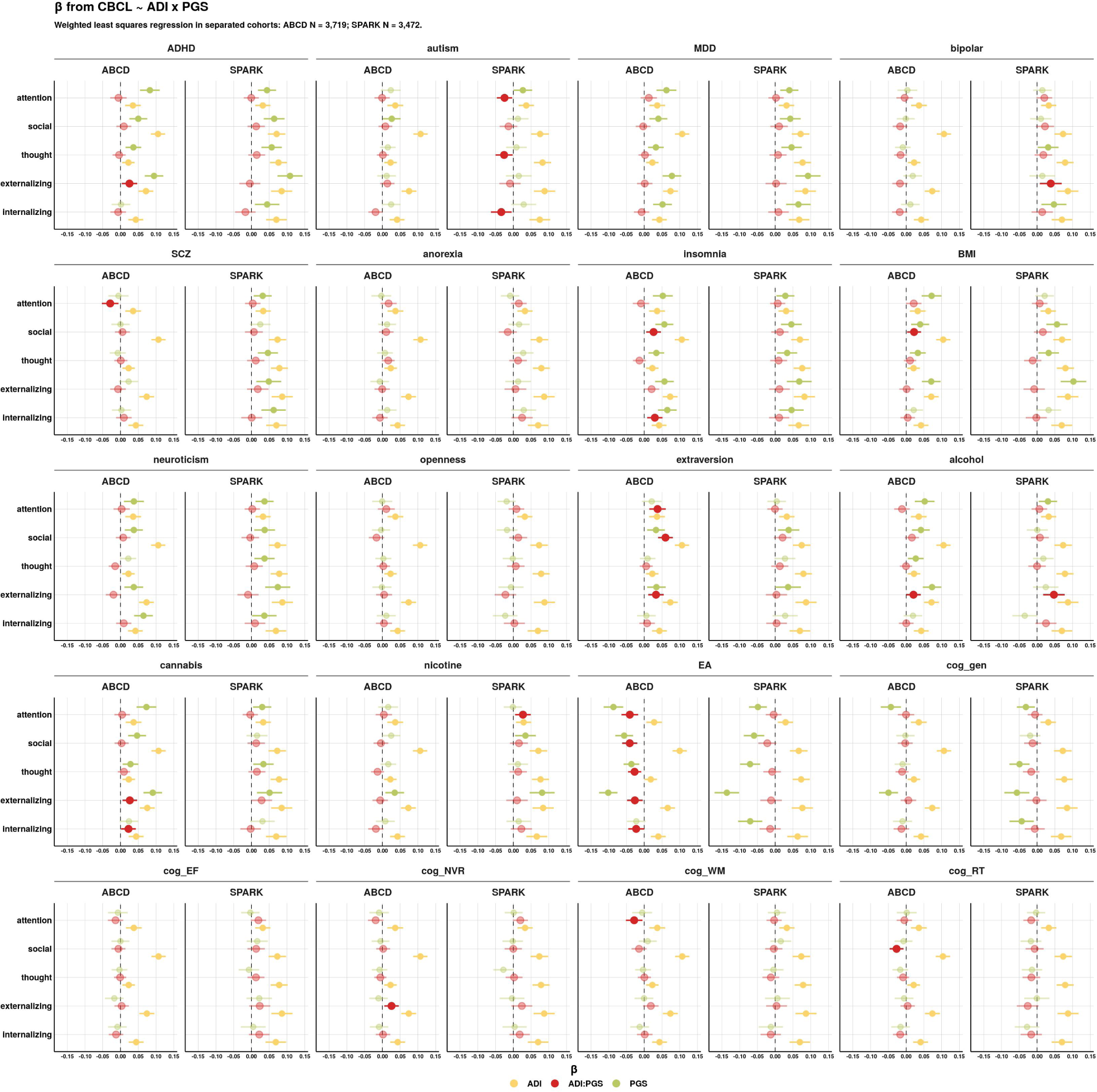
Interaction effects of the Area Deprivation Index and polygenic scores on behavioral problems calculated separately for ABCD and SPARK. The additive effects of the Area Deprivation Index (ADI), polygenic scores (PGS), and the ADI-by-PGS interaction effect were modeled using weighted linear regression: *CBCL ∼ ADI* + *PGS* + *ADI × PGS*. Prior to model input, the ADI was accounted for the 10 genetic principal components (PCs) and the 20 PGS by linear regression residualization. The *β* estimates are with the 95% confidence intervals. *β* estimates that are nominally significant (unadjusted *p <* 0.05) have a solid fill color. Models were calculated separately between the two cohorts: ABCD *N* = 3, 719 and SPARK *N* = 3, 472, with the weights re-calculated for each cohort. PGS: **MDD** = major depressive disorder, **SCZ** = schizophrenia, **alcohol** = alcohol dependency, **cannabis** = cannabis use disorder, **nicotine** = nicotine dependency, **EA** = educational attainment, **cog gen** = general cognitive performance, **cog EF** = executive functioning, **cog NVR** = non-verbal reasoning, **cog WM** = working memory, **cog RT** = reaction time.

**Figure S8.**
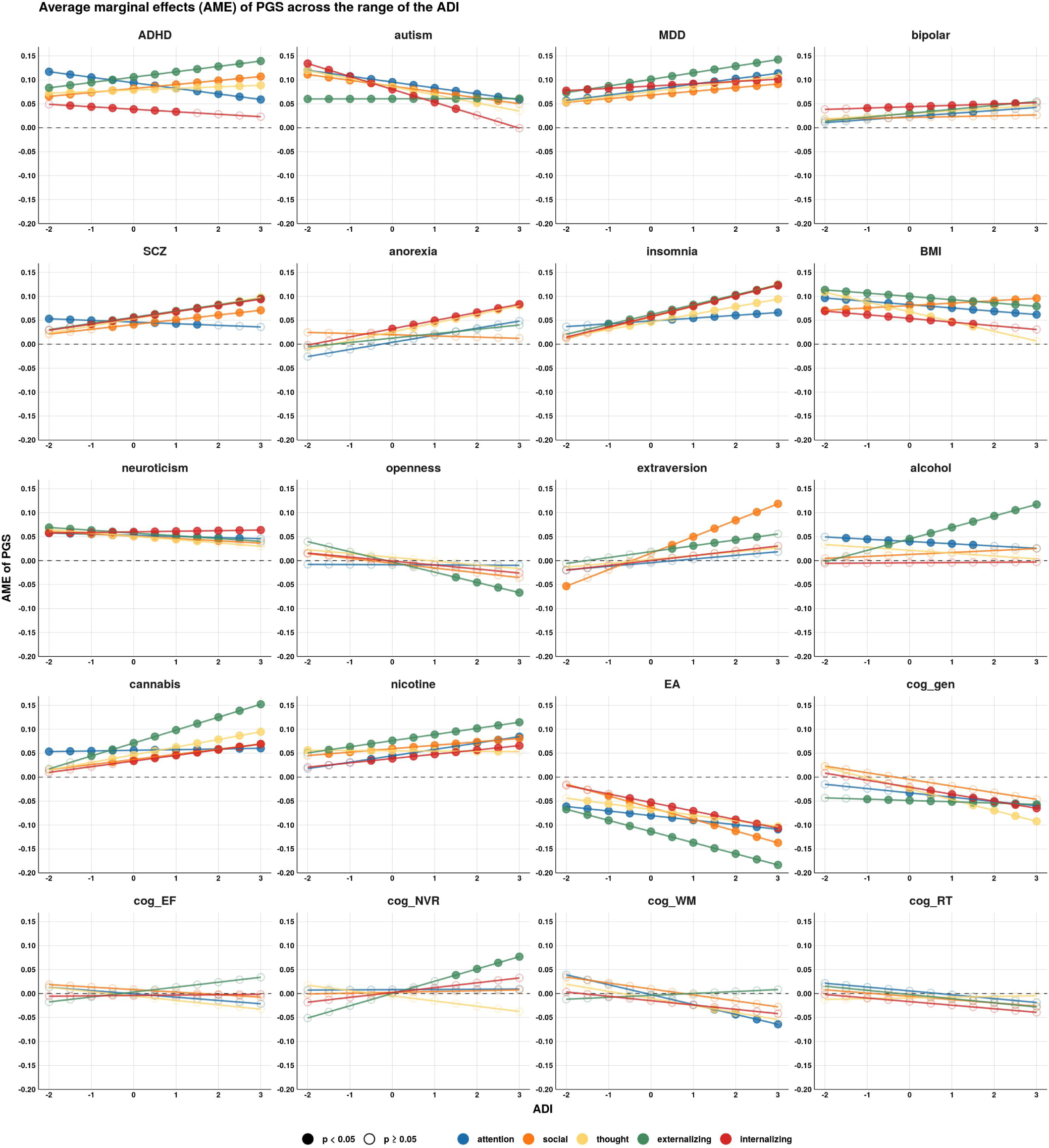
Average marginal effects of the polygenic scores across the range of the Area Deprivation Index. The Area Deprivation Index was not accounted for the 20 polygenic scores. Average marginal effects (AME) of the 20 polygenic scores (PGS) were calculated along the range of the Area Deprivation Index (ADI) at every 0.5 standard deviation from the mean using weighted least squares linear regression. The ADI was not accounted for the 20 PGS, but was residualized for the 10 genetic principal components and Z-scaled (*µ* = 0*, σ* = 1). AME that were nominally significant (unadjusted *p <* 0.05) have a solid fill color. PGS: **MDD** = major depressive disorder, **SCZ** = schizophrenia, **alcohol** = alcohol dependency, **cannabis** = cannabis use disorder, **nicotine** = nicotine dependency, **EA** = educational attainment, **cog gen** = general cognitive performance, **cog EF** = executive functioning, **cog NVR** = non-verbal reasoning, **cog WM** = working memory, **cog RT** = reaction time.

**Figure S9.**
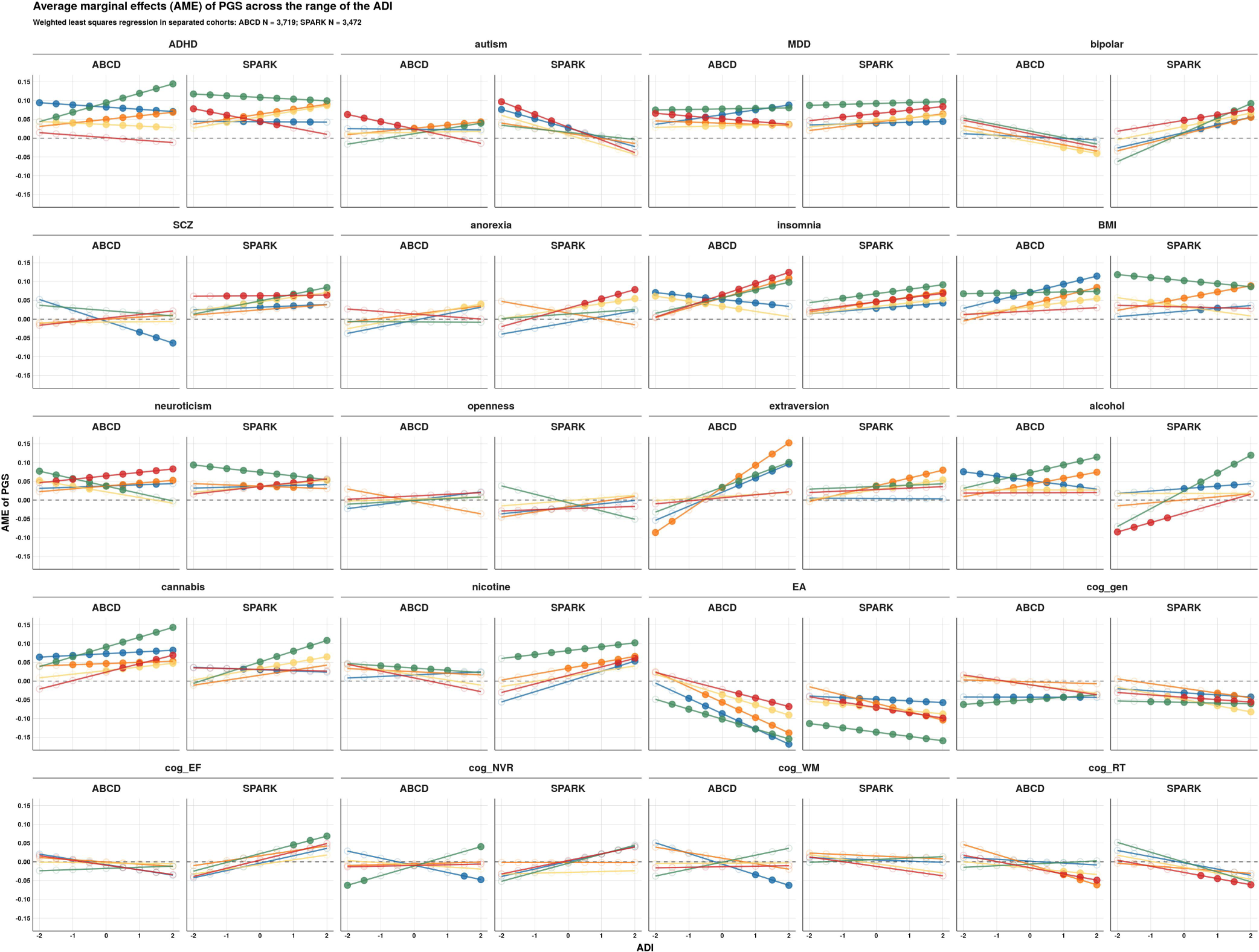
Average marginal effects of the polygenic scores across the range of the Area Deprivation Index calculated separately for ABCD and SPARK. Average marginal effects (AME) of the 20 polygenic scores (PGS) were calculated along the range of the Area Deprivation Index (ADI) at every 0.5 standard deviation from the mean using weighted least squares linear regression. The ADI was accounted for the 10 genetic principal components (PCs) and 20 PGS by linear regression residualization and then Z-scaled (*µ* = 0*, σ* = 1). AME that were nominally significant (unadjusted *p <* 0.05) have a solid fill color. PGS: **MDD** = major depressive disorder, **SCZ** = schizophrenia, **alcohol** = alcohol dependency, **cannabis** = cannabis use disorder, **nicotine** = nicotine dependency, **EA** = educational attainment, **cog gen** = general cognitive performance, **cog EF** = executive functioning, **cog NVR** = non-verbal reasoning, **cog WM** = working memory, **cog RT** = reaction time.

**Table S1.** Polygenic score associations with the Area Deprivation Index. Polygenic score (PGS) associations with the Area Deprivation Index (ADI) were performed with linear regression (*ADI ∼ PGS*). The BP statistic is the Breusch-Pagan test for heteroscedasticity (i.e., non-constant variance in the independent variable). The ADI was either: the raw value (no accounting for genetic effects), accounted for the 10 genetic principal components (PCs), or accounted for the 10 PCs and the 20 PGS. PGS: **MDD** = major depressive disorder, **SCZ** = schizophrenia, **alcohol** = alcohol dependency, **cannabis** = cannabis use disorder, **nicotine** = nicotine dependency, **EA** = educational attainment, **cog gen** = general cognitive performance, **cog EF** = executive functioning, **cog NVR** = non-verbal reasoning, **cog WM** = working memory, **cog RT** = reaction time.

| ADI acc. | PGS | weighted least squares regression |  |  |  |  |  | ordinary least squares regression |  |  |  |  |  | correlation |  |
| --- | --- | --- | --- | --- | --- | --- | --- | --- | --- | --- | --- | --- | --- | --- | --- |
| | | adj $R^2$ | pval | FDR | BP stat | BP pval | $\beta$ [95% CI] | adj $R^2$ | pval | FDR | BP stat | BP pval | $\beta$ [95% CI] | r | $\rho$ |
| PCs & PGS | ADHD | 0.00 | 0.02 | 0.09 | 0.44 | 0.51 | 0.03 [0, 0.06] | 0.00 | >.999 | >.999 | 19.29 | <.001 | 0 [-0.02, 0.02] | 0.00 | 0.00 |
| PCs | ADHD | 0.02 | <.001 | <.001 | 0.42 | 0.51 | 0.17 [0.14, 0.2] | 0.02 | <.001 | <.001 | 23.18 | <.001 | 0.14 [0.11, 0.16] | 0.14 | 0.13 |
| no acc. | ADHD | 0.02 | <.001 | <.001 | 0.11 | 0.74 | 0.17 [0.14, 0.2] | 0.02 | <.001 | <.001 | 34.13 | <.001 | 0.13 [0.11, 0.16] | 0.13 | 0.13 |
| PCs & PGS | alcohol | 0.00 | 0.58 | 0.77 | 5.16 | 0.02 | 0.01 [-0.02, 0.03] | 0.00 | >.999 | >.999 | 1.01 | 0.32 | 0 [-0.02, 0.02] | 0.00 | 0.00 |
| PCs | alcohol | 0.00 | <.001 | <.001 | 2.82 | 0.09 | 0.06 [0.04, 0.09] | 0.00 | <.001 | <.001 | 2.24 | 0.13 | 0.05 [0.03, 0.08] | 0.05 | 0.05 |
| no acc. | alcohol | 0.00 | <.001 | <.001 | 1.16 | 0.28 | 0.07 [0.04, 0.09] | 0.00 | <.001 | <.001 | 3.64 | 0.06 | 0.05 [0.03, 0.08] | 0.05 | 0.05 |
| PCs & PGS | anorexia | 0.00 | 0.30 | 0.59 | 0.68 | 0.41 | -0.01 [-0.04, 0.01] | 0.00 | >.999 | >.999 | 2.11 | 0.15 | 0 [-0.02, 0.02] | 0.00 | 0.00 |
| PCs | anorexia | 0.00 | 0.01 | 0.01 | 0.36 | 0.55 | -0.04 [-0.06, -0.01] | 0.00 | 0.04 | 0.07 | 1.12 | 0.29 | -0.02 [-0.05, 0] | -0.02 | -0.02 |
| no acc. | anorexia | 0.00 | 0.01 | 0.02 | 0.07 | 0.79 | -0.04 [-0.06, -0.01] | 0.00 | 0.05 | 0.08 | 0.47 | 0.49 | -0.02 [-0.05, 0] | -0.02 | -0.02 |
| PCs & PGS | autism | 0.00 | 0.37 | 0.68 | 0.01 | 0.93 | 0.01 [-0.01, 0.04] | 0.00 | >.999 | >.999 | 1.39 | 0.24 | 0 [-0.02, 0.02] | 0.00 | 0.00 |
| PCs | autism | 0.00 | <.001 | <.001 | 0.47 | 0.49 | 0.05 [0.02, 0.08] | 0.00 | <.001 | 0.01 | 2.94 | 0.09 | 0.03 [0.01, 0.06] | 0.03 | 0.03 |
| no acc. | autism | 0.00 | <.001 | <.001 | 0.26 | 0.61 | 0.05 [0.02, 0.07] | 0.00 | <.001 | 0.01 | 2.51 | 0.11 | 0.03 [0.01, 0.06] | 0.03 | 0.03 |
| PCs & PGS | bipolar | 0.00 | 0.54 | 0.77 | 0.01 | 0.91 | -0.01 [-0.04, 0.02] | 0.00 | >.999 | >.999 | 0.15 | 0.69 | 0 [-0.02, 0.02] | 0.00 | 0.00 |
| PCs | bipolar | 0.00 | 0.52 | 0.62 | 0.14 | 0.71 | -0.01 [-0.04, 0.02] | 0.00 | 0.89 | 0.93 | 0.13 | 0.72 | 0 [-0.02, 0.02] | 0.00 | 0.00 |
| no acc. | bipolar | 0.00 | 0.59 | 0.65 | 0.34 | 0.56 | -0.01 [-0.03, 0.02] | 0.00 | 0.89 | 0.94 | 0.05 | 0.83 | 0 [-0.02, 0.02] | 0.00 | 0.00 |
| PCs & PGS | BMI | 0.00 | <.001 | <.001 | 1.26 | 0.26 | 0.05 [0.03, 0.08] | 0.00 | >.999 | >.999 | 11.54 | <.001 | 0 [-0.02, 0.02] | 0.00 | 0.00 |
| PCs | BMI | 0.02 | <.001 | <.001 | 0.83 | 0.36 | 0.17 [0.15, 0.2] | 0.01 | <.001 | <.001 | 14.33 | <.001 | 0.12 [0.1, 0.14] | 0.12 | 0.11 |
| no acc. | BMI | 0.02 | <.001 | <.001 | 2.62 | 0.11 | 0.17 [0.14, 0.2] | 0.01 | <.001 | <.001 | 22.81 | <.001 | 0.12 [0.09, 0.14] | 0.12 | 0.11 |
| PCs & PGS | cannabis | 0.00 | 0.60 | 0.77 | 1.81 | 0.18 | 0.01 [-0.02, 0.03] | 0.00 | >.999 | >.999 | 1.81 | 0.18 | 0 [-0.02, 0.02] | 0.00 | 0.00 |
| PCs | cannabis | 0.00 | <.001 | <.001 | 1.07 | 0.30 | 0.07 [0.04, 0.09] | 0.00 | <.001 | <.001 | 4.84 | 0.03 | 0.06 [0.04, 0.08] | 0.06 | 0.06 |
| no acc. | cannabis | 0.00 | <.001 | <.001 | 1.34 | 0.25 | 0.07 [0.04, 0.09] | 0.00 | <.001 | <.001 | 4.61 | 0.03 | 0.06 [0.03, 0.08] | 0.06 | 0.06 |
| PCs & PGS | cog EF | 0.00 | 0.17 | 0.38 | 5.38 | 0.02 | 0.02 [-0.01, 0.04] | 0.00 | >.999 | >.999 | 0.85 | 0.36 | 0 [-0.02, 0.02] | 0.00 | 0.00 |
| PCs | cog EF | 0.00 | 0.04 | 0.05 | 2.08 | 0.15 | 0.03 [0, 0.06] | 0.00 | 0.33 | 0.42 | 0.17 | 0.68 | 0.01 [-0.01, 0.03] | 0.01 | 0.01 |
| no acc. | cog EF | 0.00 | 0.04 | 0.06 | 1.69 | 0.19 | 0.03 [0, 0.06] | 0.00 | 0.35 | 0.44 | 0.07 | 0.79 | 0.01 [-0.01, 0.03] | 0.01 | 0.01 |
| PCs & PGS | cog gen | 0.00 | 0.09 | 0.24 | 0.40 | 0.53 | -0.02 [-0.05, 0] | 0.00 | >.999 | >.999 | 3.15 | 0.08 | 0 [-0.02, 0.02] | 0.00 | 0.00 |
| PCs | cog gen | 0.01 | <.001 | <.001 | 0.87 | 0.35 | -0.1 [-0.13, -0.07] | 0.01 | <.001 | <.001 | 3.57 | 0.06 | -0.08 [-0.1, -0.05] | -0.08 | -0.07 |
| no acc. | cog gen | 0.01 | <.001 | <.001 | 0.49 | 0.48 | -0.1 [-0.13, -0.07] | 0.01 | <.001 | <.001 | 6.80 | 0.01 | -0.07 [-0.1, -0.05] | -0.07 | -0.07 |
| PCs & PGS | cog NVR | 0.00 | 0.71 | 0.84 | 0.59 | 0.44 | -0.01 [-0.03, 0.02] | 0.00 | >.999 | >.999 | 1.21 | 0.27 | 0 [-0.02, 0.02] | 0.00 | 0.01 |
| PCs | cog NVR | 0.00 | 0.02 | 0.04 | 0.89 | 0.35 | -0.03 [-0.06, 0] | 0.00 | 0.03 | 0.04 | 1.91 | 0.17 | -0.03 [-0.05, 0] | -0.03 | -0.02 |
| no acc. | cog NVR | 0.00 | 0.03 | 0.05 | 1.34 | 0.25 | -0.03 [-0.06, 0] | 0.00 | 0.03 | 0.05 | 3.07 | 0.08 | -0.03 [-0.05, 0] | -0.03 | -0.02 |
| PCs & PGS | cog RT | 0.00 | 0.79 | 0.88 | 2.49 | 0.11 | 0 [-0.03, 0.02] | 0.00 | >.999 | >.999 | 0.15 | 0.70 | 0 [-0.02, 0.02] | 0.00 | 0.00 |
| PCs | cog RT | 0.00 | 0.63 | 0.70 | 1.49 | 0.22 | 0.01 [-0.02, 0.03] | 0.00 | 0.47 | 0.55 | 0.13 | 0.72 | 0.01 [-0.01, 0.03] | 0.01 | 0.01 |
| no acc. | cog RT | 0.00 | 0.52 | 0.61 | 0.77 | 0.38 | 0.01 [-0.02, 0.04] | 0.00 | 0.48 | 0.57 | 0.23 | 0.63 | 0.01 [-0.01, 0.03] | 0.01 | 0.01 |
| PCs & PGS | cog WM | 0.00 | 0.99 | 0.99 | 0.78 | 0.38 | 0 [-0.03, 0.03] | 0.00 | >.999 | >.999 | 0.32 | 0.57 | 0 [-0.02, 0.02] | 0.00 | 0.00 |
| PCs | cog WM | 0.00 | 0.91 | 0.91 | 0.68 | 0.41 | 0 [-0.03, 0.03] | 0.00 | 0.95 | 0.95 | 0.39 | 0.53 | 0 [-0.02, 0.02] | 0.00 | 0.00 |
| no acc. | cog WM | 0.00 | 0.94 | 0.94 | 2.61 | 0.11 | 0 [-0.03, 0.03] | 0.00 | 0.95 | 0.95 | 0.71 | 0.40 | 0 [-0.02, 0.02] | 0.00 | 0.00 |
| PCs & PGS | EA | 0.00 | <.001 | <.001 | 1.28 | 0.26 | -0.08 [-0.1, -0.05] | 0.00 | >.999 | >.999 | 55.61 | <.001 | 0 [-0.02, 0.02] | 0.00 | 0.00 |
| PCs | EA | 0.08 | <.001 | <.001 | 1.84 | 0.18 | -0.32 [-0.34, -0.29] | 0.06 | <.001 | <.001 | 59.97 | <.001 | -0.24 [-0.27, -0.22] | -0.24 | -0.24 |
| no acc. | EA | 0.07 | <.001 | <.001 | 2.09 | 0.15 | -0.31 [-0.34, -0.29] | 0.06 | <.001 | <.001 | 68.34 | <.001 | -0.24 [-0.26, -0.21] | -0.24 | -0.23 |
| PCs & PGS | extraversion | 0.00 | 0.62 | 0.77 | 1.77 | 0.18 | -0.01 [-0.03, 0.02] | 0.00 | >.999 | >.999 | 1.70 | 0.19 | 0 [-0.02, 0.02] | 0.00 | 0.00 |
| PCs | extraversion | 0.00 | 0.12 | 0.16 | 1.41 | 0.23 | -0.02 [-0.05, 0.01] | 0.00 | 0.23 | 0.33 | 2.37 | 0.12 | -0.01 [-0.04, 0.01] | -0.01 | -0.02 |
| no acc. | extraversion | 0.00 | 0.12 | 0.16 | 0.71 | 0.40 | -0.02 [-0.05, 0.01] | 0.00 | 0.25 | 0.35 | 1.47 | 0.23 | -0.01 [-0.04, 0.01] | -0.01 | -0.02 |
| PCs & PGS | insomnia | 0.00 | <.001 | 0.02 | 3.39 | 0.07 | 0.04 [0.01, 0.06] | 0.00 | >.999 | >.999 | 15.45 | <.001 | 0 [-0.02, 0.02] | 0.00 | -0.01 |
| PCs | insomnia | 0.01 | <.001 | <.001 | 1.06 | 0.30 | 0.11 [0.09, 0.14] | 0.01 | <.001 | <.001 | 14.37 | <.001 | 0.08 [0.05, 0.1] | 0.08 | 0.06 |
| no acc. | insomnia | 0.01 | <.001 | <.001 | 4.93 | 0.03 | 0.11 [0.09, 0.14] | 0.01 | <.001 | <.001 | 20.94 | <.001 | 0.07 [0.05, 0.1] | 0.07 | 0.06 |
| PCs & PGS | MDD | 0.00 | 0.04 | 0.13 | 0.06 | 0.80 | 0.03 [0, 0.05] | 0.00 | >.999 | >.999 | 7.66 | 0.01 | 0 [-0.02, 0.02] | 0.00 | 0.00 |
| PCs | MDD | 0.01 | <.001 | <.001 | 0.04 | 0.84 | 0.13 [0.1, 0.16] | 0.01 | <.001 | <.001 | 9.99 | <.001 | 0.1 [0.08, 0.12] | 0.10 | 0.09 |
| no acc. | MDD | 0.01 | <.001 | <.001 | 0.00 | 0.99 | 0.13 [0.1, 0.15] | 0.01 | <.001 | <.001 | 11.93 | <.001 | 0.1 [0.07, 0.12] | 0.10 | 0.09 |
| PCs & PGS | neuroticism | 0.00 | 0.07 | 0.21 | 0.50 | 0.48 | 0.02 [0, 0.05] | 0.00 | >.999 | >.999 | 4.05 | 0.04 | 0 [-0.02, 0.02] | 0.00 | 0.00 |
| PCs | neuroticism | 0.01 | <.001 | <.001 | 1.66 | 0.20 | 0.1 [0.07, 0.12] | 0.01 | <.001 | <.001 | 3.89 | 0.05 | 0.07 [0.05, 0.1] | 0.07 | 0.07 |
| no acc. | neuroticism | 0.01 | <.001 | <.001 | 0.69 | 0.41 | 0.1 [0.07, 0.12] | 0.01 | <.001 | <.001 | 5.19 | 0.02 | 0.07 [0.05, 0.1] | 0.07 | 0.07 |
| PCs & PGS | nicotine | 0.00 | <.001 | 0.02 | 0.95 | 0.33 | 0.04 [0.01, 0.06] | 0.00 | >.999 | >.999 | 16.01 | <.001 | 0 [-0.02, 0.02] | 0.00 | 0.00 |
| PCs | nicotine | 0.01 | <.001 | <.001 | 0.89 | 0.35 | 0.14 [0.12, 0.17] | 0.01 | <.001 | <.001 | 16.28 | <.001 | 0.1 [0.08, 0.13] | 0.10 | 0.10 |
| no acc. | nicotine | 0.01 | <.001 | <.001 | 2.23 | 0.14 | 0.14 [0.12, 0.17] | 0.01 | <.001 | <.001 | 22.42 | <.001 | 0.1 [0.08, 0.12] | 0.10 | 0.09 |
| PCs & PGS | openness | 0.00 | 0.44 | 0.74 | 1.14 | 0.29 | 0.01 [-0.02, 0.04] | 0.00 | >.999 | >.999 | 0.00 | 0.97 | 0 [-0.02, 0.02] | 0.00 | -0.01 |
| PCs | openness | 0.00 | 0.69 | 0.73 | 3.65 | 0.06 | 0.01 [-0.02, 0.03] | 0.00 | 0.59 | 0.65 | 0.31 | 0.58 | -0.01 [-0.03, 0.02] | -0.01 | -0.01 |
| no acc. | openness | 0.00 | 0.70 | 0.73 | 5.02 | 0.03 | 0.01 [-0.02, 0.03] | 0.00 | 0.60 | 0.67 | 0.29 | 0.59 | -0.01 [-0.03, 0.02] | -0.01 | -0.01 |
| PCs & PGS | SCZ | 0.00 | 0.85 | 0.89 | 0.42 | 0.52 | 0 [-0.03, 0.02] | 0.00 | >.999 | >.999 | 2.16 | 0.14 | 0 [-0.02, 0.02] | 0.00 | 0.00 |
| PCs | SCZ | 0.00 | 0.48 | 0.60 | 0.02 | 0.90 | 0.01 [-0.02, 0.04] | 0.00 | 0.30 | 0.40 | 2.13 | 0.14 | 0.01 [-0.01, 0.04] | 0.01 | 0.01 |
| no acc. | SCZ | 0.00 | 0.48 | 0.61 | 0.01 | 0.94 | 0.01 [-0.02, 0.04] | 0.00 | 0.31 | 0.42 | 2.02 | 0.16 | 0.01 [-0.01, 0.04] | 0.01 | 0.01 |

**Table S2.** Area Deprivation Index associations with behavioral problems. Area Deprivation Index (ADI) associations with the Child Behavior Checklist (CBCL) subscale scores were performed with linear regression (*CBCL ∼ ADI*). The BP statistic is the Breusch-Pagan test for heteroscedasticity (i.e., non-constant variance in the independent variable). The ADI was either: the raw value (no accounting for genetic effects), accounted for the 10 genetic principal components (PCs), or accounted for the 10 PCs and the 20 PGS.

| CBCL | ADI acc. | weighted least squares regression |  |  |  |  |  | ordinary least squares regression |  |  |  |  |  | correlation |  |
| --- | --- | --- | --- | --- | --- | --- | --- | --- | --- | --- | --- | --- | --- | --- | --- |
| | | adj $R^2$ | pval | FDR | BP stat | BP pval | $\beta$ [95% CI] | adj $R^2$ | pval | FDR | BP stat | BP pval | $\beta$ [95% CI] | $r$ | $\rho$ |
| attention | no acc. | 0.06 | <.001 | <.001 | 15.36 | <.001 | 0.21 [0.19, 0.23] | 0.05 | <.001 | <.001 | 8.68 | <.001 | 0.23 [0.2, 0.25] | 0.23 | 0.23 |
| attention | PCs | 0.05 | <.001 | <.001 | 9.34 | <.001 | 0.19 [0.17, 0.21] | 0.04 | <.001 | <.001 | 5.75 | 0.02 | 0.21 [0.19, 0.23] | 0.21 | 0.21 |
| attention | PCs & PGS | 0.04 | <.001 | <.001 | 4.47 | 0.03 | 0.16 [0.14, 0.18] | 0.03 | <.001 | <.001 | 2.32 | 0.13 | 0.17 [0.14, 0.19] | 0.17 | 0.17 |
| social | no acc. | 0.08 | <.001 | <.001 | 6.86 | 0.01 | 0.25 [0.23, 0.27] | 0.07 | <.001 | <.001 | 11.84 | <.001 | 0.26 [0.24, 0.28] | 0.26 | 0.26 |
| social | PCs | 0.07 | <.001 | <.001 | 8.75 | <.001 | 0.24 [0.22, 0.25] | 0.06 | <.001 | <.001 | 12.00 | <.001 | 0.24 [0.22, 0.27] | 0.24 | 0.24 |
| social | PCs & PGS | 0.06 | <.001 | <.001 | 6.19 | 0.01 | 0.21 [0.19, 0.23] | 0.04 | <.001 | <.001 | 7.44 | 0.01 | 0.21 [0.18, 0.23] | 0.21 | 0.21 |
| thought | no acc. | 0.07 | <.001 | <.001 | 45.05 | <.001 | 0.23 [0.21, 0.25] | 0.07 | <.001 | <.001 | 44.53 | <.001 | 0.26 [0.23, 0.28] | 0.26 | 0.26 |
| thought | PCs | 0.06 | <.001 | <.001 | 49.18 | <.001 | 0.22 [0.2, 0.24] | 0.06 | <.001 | <.001 | 45.10 | <.001 | 0.24 [0.21, 0.26] | 0.24 | 0.24 |
| thought | PCs & PGS | 0.05 | <.001 | <.001 | 42.29 | <.001 | 0.19 [0.17, 0.21] | 0.04 | <.001 | <.001 | 34.09 | <.001 | 0.2 [0.18, 0.22] | 0.20 | 0.20 |
| externalizing | no acc. | 0.06 | <.001 | <.001 | 51.63 | <.001 | 0.22 [0.2, 0.24] | 0.05 | <.001 | <.001 | 54.92 | <.001 | 0.22 [0.2, 0.25] | 0.22 | 0.22 |
| externalizing | PCs | 0.06 | <.001 | <.001 | 55.43 | <.001 | 0.21 [0.19, 0.23] | 0.04 | <.001 | <.001 | 56.39 | <.001 | 0.21 [0.19, 0.23] | 0.21 | 0.21 |
| externalizing | PCs & PGS | 0.04 | <.001 | <.001 | 31.75 | <.001 | 0.17 [0.15, 0.19] | 0.03 | <.001 | <.001 | 28.85 | <.001 | 0.16 [0.14, 0.19] | 0.16 | 0.16 |
| internalizing | no acc. | 0.04 | <.001 | <.001 | 30.04 | <.001 | 0.18 [0.16, 0.2] | 0.04 | <.001 | <.001 | 36.64 | <.001 | 0.2 [0.18, 0.22] | 0.20 | 0.20 |
| internalizing | PCs | 0.04 | <.001 | <.001 | 26.34 | <.001 | 0.17 [0.15, 0.19] | 0.03 | <.001 | <.001 | 30.37 | <.001 | 0.18 [0.16, 0.2] | 0.18 | 0.19 |
| internalizing | PCs & PGS | 0.03 | <.001 | <.001 | 17.03 | <.001 | 0.15 [0.13, 0.17] | 0.02 | <.001 | <.001 | 17.94 | <.001 | 0.15 [0.13, 0.18] | 0.15 | 0.16 |

**Table S3.**
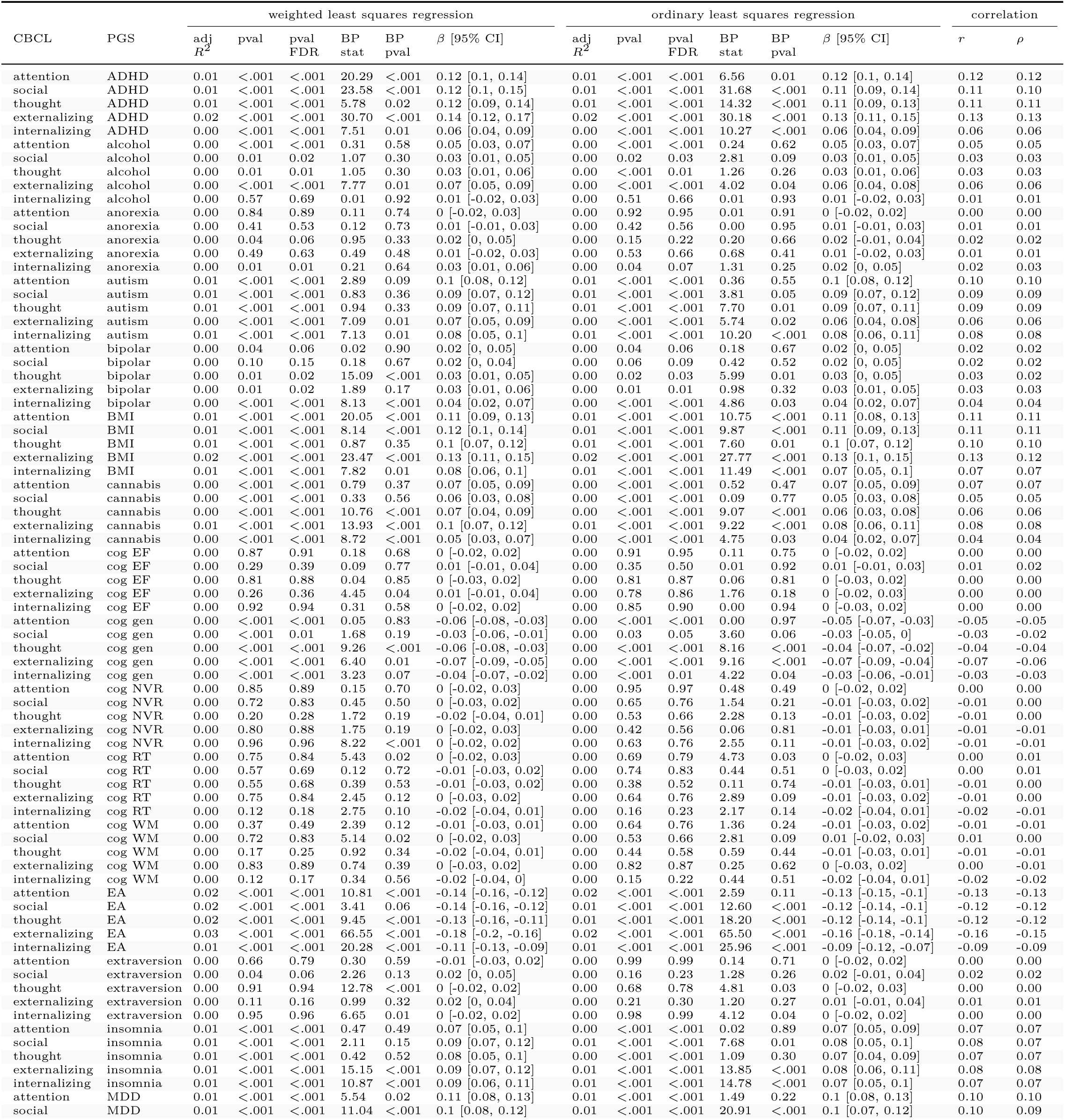
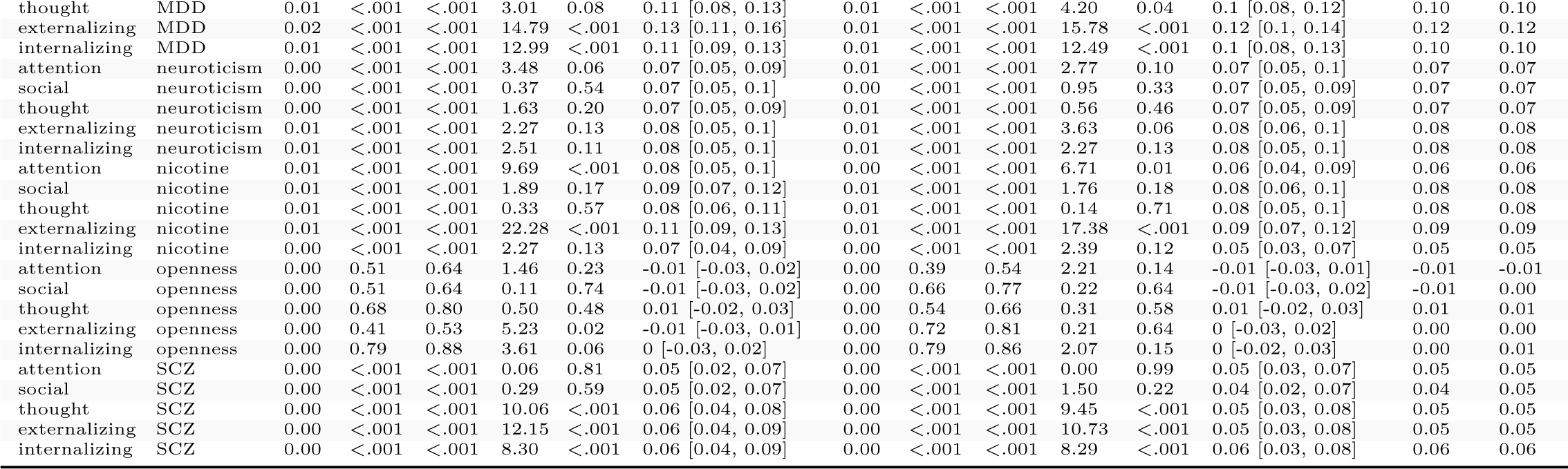
Polygenic score associations with behavioral problems. Polygenic score (PGS) associations with the Child Behavior Checklist (CBCL) subscale scores were performed with linear regression (*CBCL ∼ PGS*). The BP statistic is the Breusch-Pagan test for heteroscedasticity (i.e., non-constant variance in the independent variable). PGS: **MDD** = major depressive disorder, **SCZ** = schizophrenia, **alcohol** = alcohol dependency, **cannabis** = cannabis use disorder, **nicotine** = nicotine dependency, **EA** = educational attainment, **cog gen** = general cognitive performance, **cog EF** = executive functioning, **cog NVR** = non-verbal reasoning, **cog WM** = working memory, **cog RT** = reaction time.

**Table S4.**
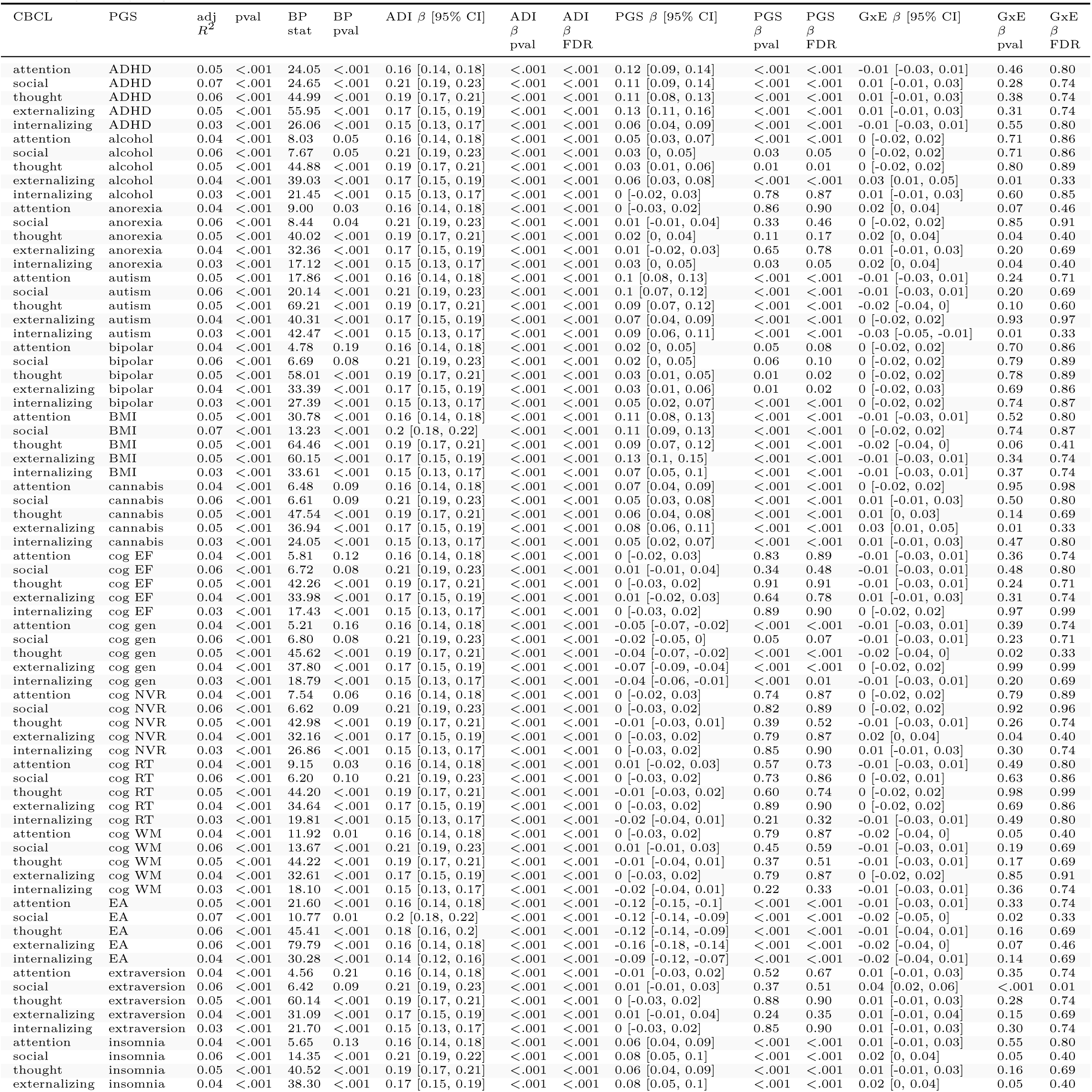
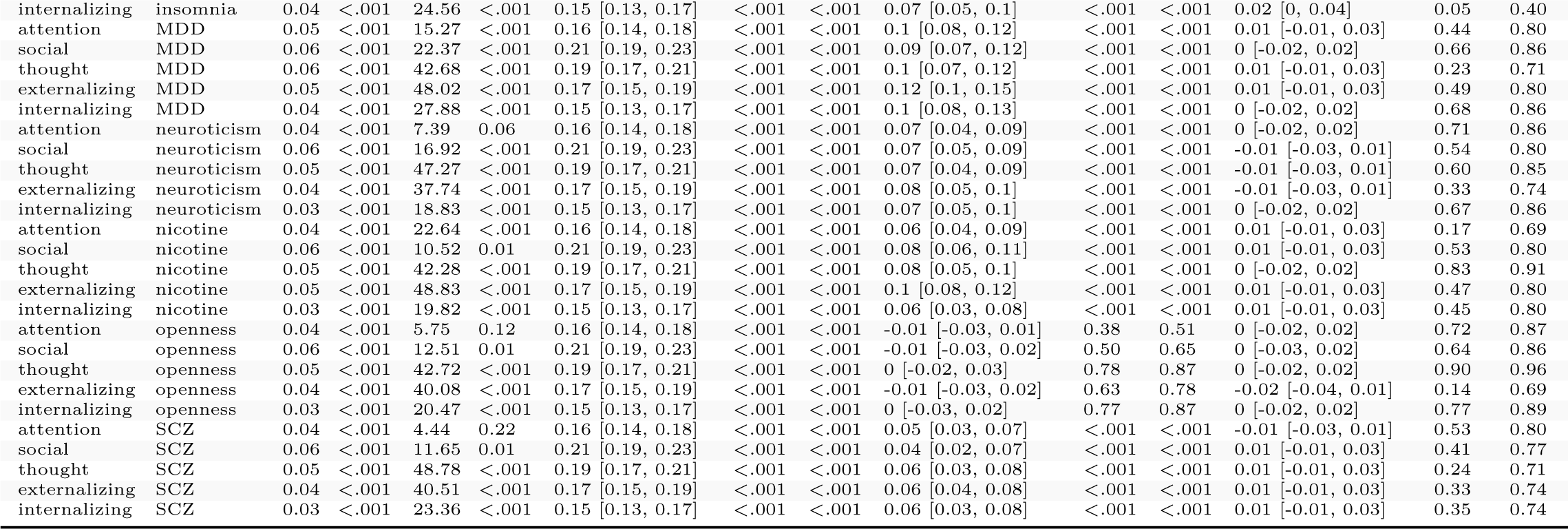
Interaction effects of the Area Deprivation Index and polygenic scores on behavioral problems. The independent effects of the Area Deprivation Index (ADI), polygenic scores (PGS), and the ADI-by-PGS interaction (GxE) effects were modeled using weighted linear regression: *CBCL ∼ ADI* + *PGS* + *ADI × PGS*. The ADI was accounted for the 20 PGS prior to model input. The BP statistic is the Breusch-Pagan test for heteroscedasticity (i.e., non-constant variance in the independent variable). PGS: **MDD** = major depressive disorder, **SCZ** = schizophrenia, **alcohol** = alcohol dependency, **cannabis** = cannabis use disorder, **nicotine** = nicotine dependency, **EA** = educational attainment, **cog gen** = general cognitive performance, **cog EF** = executive functioning, **cog NVR** = non-verbal reasoning, **cog WM** = working memory, **cog RT** = reaction time.

**Table S5.**
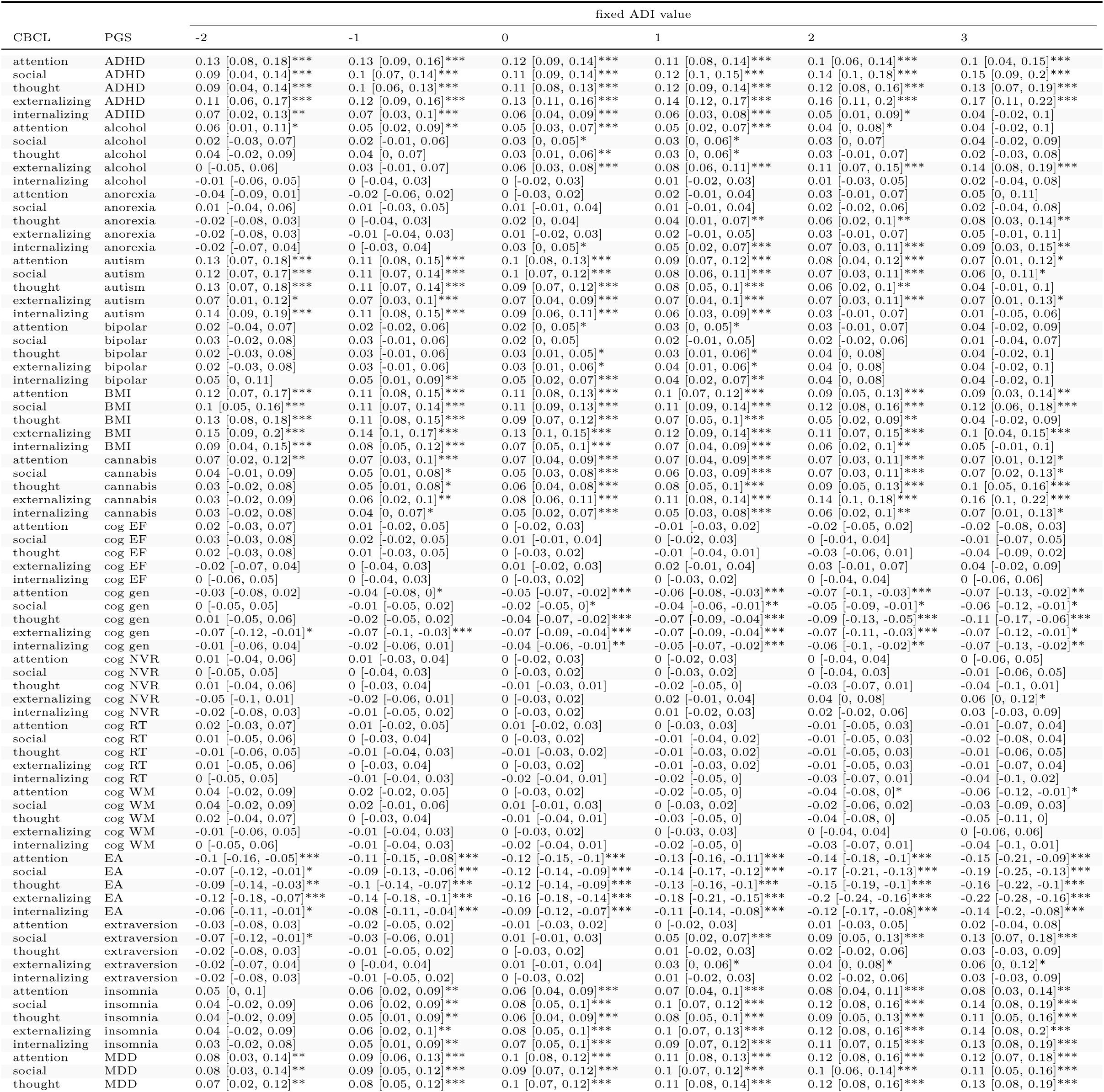
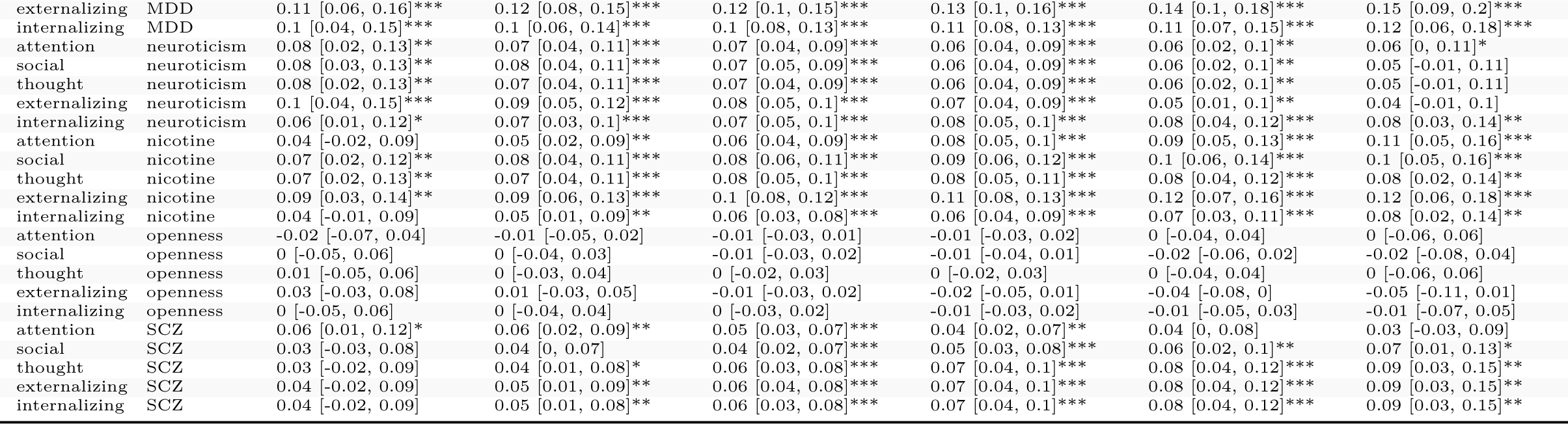
Average marginal effects of polygenic scores across the range of the Area Deprivation Index. Average marginal effects (AME) of the 20 polygenic scores (PGS) were calculated along the Area Deprivation Index (ADI) at every 0.5 value of the ADI using weighted linear regression. The ADI was accounted for the 20 PGS prior to calculating the AME. Nominal significance of AME indicated by asterisks: *∗* = *p <* 0.05, ** = p < 0.01*, ∗∗∗* = *p <* 0.001. PGS: **MDD** = major depressive disorder, **SCZ** = schizophrenia, **alcohol** = alcohol dependency, **cannabis** = cannabis use disorder, **nicotine** = nicotine dependency, **EA** = educational attainment, **cog gen** = general cognitive performance, **cog EF** = executive functioning, **cog NVR** = non-verbal reasoning, **cog WM** = working memory, **cog RT** = reaction time.

